# Abatacept for Treatment of Adults Hospitalized with Moderate or Severe Covid-19

**DOI:** 10.1101/2022.09.22.22280247

**Authors:** Emily R. Ko, Kevin J. Anstrom, Reynold A. Panettieri, Anne M. Lachiewicz, Martin Maillo, Jane A. O’Halloran, Cynthia Boucher, P. Brian Smith, Matthew W. McCarthy, Patricia Segura Nunez, Sabina Mendivil Tuchia de Tai, Akram Khan, Alfredo J. Mena Lora, Matthias Salathe, Eyal Kedar, Gerardo Capo, Daniel Rodríguez Gonzalez, Thomas F. Patterson, Christopher Palma, Horacio Ariza, Maria Patelli Lima, John Blamoun, Esteban C. Nannini, Eduardo Sprinz, Analia Mykietiuk, Jennifer P. Wang, Luis Parra-Rodriguez, Tatyana Der, Kate Willsey, Daniel K. Benjamin, Jun Wen, Pearl Zakroysky, Susan Halabi, Adam Silverstein, Steven E. McNulty, Sean M. O’Brien, Hussein R. Al-Khalidi, Sandra Butler, Jane Atkinson, Stacey J. Adam, Soju Chang, Michael A. Maldonado, Michael Proscham, Lisa LaVange, Samuel A. Bozzette, William G. Powderly, the ACTIV-1 IM study group members

**Affiliations:** Duke University Medical Center, Durham, NC; University of North Carolina, Chapel Hill, NC; Robert Wood Johnson Medical School, School, New Brunswick, NJ; Sanatorio Diagnostico, Diagnostico, Santa Fe, Argentina; Washington University St. Louis, St Louis, MO; National Center for Advancing Translational Sciences; Duke Clinical Research Institute (DCRI); Weill Cornell Medicine in New York; Hospital Nacional Hipolito Unanue, Lima, Peru; Hospital Central de la Fuerza Aerea del Peru, Lima, Peru; Oregon Health and Science University, Portland, OR; University of Illinois at Chicago, Chicago, IL; University of Kansas Medical Center, Kansas City, KS; St. Lawrence Health, Potsdam, NY; Trinitas Hospital, Elizabeth, NJ; Nuevo Hospital Civil de Guadalajara Juan I. Menchaca, Guadalajara, Mexico; University of Texas Health Science Center at San Antonio, San Antonio, TX; University of Rochester Medical School Medicine and Dentistry, Rochester, NY; Clinica Central S.A., Villa Regina, Argentina; Hospital e Maternidade Celso Pierro - PUC Campinas; MidMichigan Medical Center – Midland, Midland, MI; Sanatorio Britanico. Santa Fe, Argentina; Hospital de Clinicas de Porto Alegre HCPA, Porto Alegre, Brazil; Instituto Medico Platense, La Plata, Argentina; University of Massachusetts Medical Center, Worcester, MA; Washington University St. Louis; Technical Resources International (TRI), Bethesda MD; Foundation for the National Institutes of Health (FNIH); Bristol Myers Squibb; National Institute of Allergy and Infectious Diseases; Washington University in St Louis School of Medicine

**Author notes:** **Address for correspondence:** William G. Powderly, MD, Division of Infectious Diseases, Department of Medicine, Washington University School of Medicine, St. Louis, MO 63110.

**Keywords:** abatacept, immune checkpoint inhibitors, Covid-19, therapeutics, SARS-CoV-2, T-Lymphocyte, Regulatory

## Abstract

**Background:** We investigated whether abatacept, a selective costimulation modulator, provides additional benefit when added to standard-of-care for patients hospitalized with Covid-19.

**Methods:** We conducted a master protocol to investigate immunomodulators for potential benefit treating patients hospitalized with Covid-19 and report results for abatacept. Intravenous abatacept (one-time dose 10 mg/kg, maximum dose 1000 mg) plus standard of care (SOC) was compared with shared placebo plus SOC. Primary outcome was time-to-recovery by day 28. Key secondary endpoints included 28-day mortality.

**Results:** Between October 16, 2020 and December 31, 2021, a total of 1019 participants received study treatment (509 abatacept; 510 shared placebo), constituting the modified intention-to-treat cohort. Participants had a mean age 54.8 (SD 14.6) years, 60.5% were male, 44.2% Hispanic/Latino and 13.7% Black. No statistically significant difference for the primary endpoint of time-to-recovery was found with a recovery-rate-ratio of 1.14 (95% CI 1.00–1.29; p=0.057) compared with placebo. We observed a substantial improvement in 28-day all-cause mortality with abatacept versus placebo (11.0% vs. 15.1%; odds ratio [OR] 0.62 [95% CI 0.41– 0.94]), leading to 38% lower odds of dying. Improvement in mortality occurred for participants requiring oxygen/noninvasive ventilation at randomization. Subgroup analysis identified the strongest effect in those with baseline C-reactive protein >75mg/L. We found no statistically significant differences in adverse events, with safety composite index slightly favoring abatacept. Rates of secondary infections were similar (16.1% for abatacept; 14.3% for placebo).

**Conclusions:** Addition of single-dose intravenous abatacept to standard-of-care demonstrated no statistically significant change in time-to-recovery, but improved 28-day mortality.

**Trial registration:** ClinicalTrials.gov (NCT04593940).

## INTRODUCTION

A hyperactive immune response, known as cytokine storm, is a hallmark of severe Covid-19.^1^ The RECOVERY trial found dexamethasone had a mortality benefit for patients with Covid-19 on oxygen,^2^ and became a mainstay anti-inflammatory treatment for patients requiring oxygen. However, substantial morbidity and mortality remained. Several immunomodulators were explored as Covid-19 therapeutics since these agents can neutralize destructive cytokine signaling and restore immunoregulatory cascades.^1^ Abatacept, a fusion protein of the extracellular portion of the T-lymphocyte CTLA-4 domain and a fragment of the Fc portion of human immunoglobin, mimics complex inhibition of T-cell costimulatory signaling to restore homeostasis to the immune system.^3^ Abatacept offers significant therapeutic benefits to patients with autoimmune and hyperinflammatory illnesses.^4,5^ Preclinical data in murine infection models showed that abatacept limited influenza- and sepsis-induced inflammatory cascades that cause tissue damage and preserved viral clearance in the influenza model.^6-9^ Abatacept’s unique function could balance pathogen clearance and immune tolerance.^10^ The only data currently available for abatacept in patients with Covid-19 are longitudinal observation studies evaluating severity of Covid-19 in patients on disease-modifying anti-rheumatic drugs (DMARDs). These studies suggest patients taking abatacept for rheumatologic disorders do not have a higher risk of hospitalization and severe symptoms than those on other DMARDs.^11-14^

In April 2020, the National Institutes of Health (NIH) launched a public-private collaboration, Accelerating Covid-19 Therapeutic Interventions and Vaccines (ACTIV), to develop a coordinated research response. The ACTIV-1 Immune Modulators (IM) master protocol evaluated multiple immunomodulatory agents for improved outcomes in participants hospitalized with moderate-to-severe Covid-19. We selected abatacept for inclusion based on its novel mechanism, significant efficacy, and safety profile in participants with inflammatory disorders. We report the results of abatacept plus standard of care compared with placebo plus standard of care in patients with Covid-19.

## METHODS

### Study design

ACTIV-1 IM, a randomized, double-blind, placebo-controlled master protocol^15^ evaluated three immune modulators plus standard of care compared with shared placebo plus standard of care in separate arms. An innovative design feature is utilization of a shared placebo, minimizing the number of participants receiving placebo and reducing the sample size needed to retain adequate power to evaluate multiple agents (**Supplementary Appendix**). The protocol and statistical analysis plan are available in a **Supplementary Appendix**.^15^

### Eligibility

The protocol was approved by relevant review boards; all patients provided informed consent. Participants ≥18 years of age with confirmed SARS-CoV-2 infection of ≤14 days, anticipated hospital stay of ≥72 hours, and evidence of pulmonary involvement were eligible. Candidates were excluded with pregnancy, liver function test >10x normal, chronic liver disease, acute kidney injury with glomerular filtration rate<30 (stable chronic renal insufficiency permitted), severe heart failure, severe neutropenia or lymphopenia, known untreated infection other than Covid-19, or those who received cytotoxic or biologic targeted immunomodulators within 4 weeks or 5 half-lives before screening.

### Procedures

Abatacept was administered on day 1 as a single-dose intravenous infusion of 10 mg/kg (maximum dose 1000 mg) or matching placebo saline infusion. Participants received standard supportive care at the site hospital, including remdesivir (study provided [Gilead Sciences, Foster City, CA]), dexamethasone, or emergency-use-authorized (EUA) monoclonal antibodies per national guidelines. Clinical status and safety data were captured daily during hospitalization through day 28 and post-discharge at days 8, 11, 15, 29, and 60.

### Endpoints

The primary outcome of time to recovery at day 28 was evaluated using an 8-point ordinal scale (OS) (**Table 1**). Recovery was identified as the first day a participant attained category 6, 7, or 8. Key secondary endpoints were pre-specified as 28-day mortality and clinical status at day 14 assessed by OS improvement from baseline.

**Table 1.**
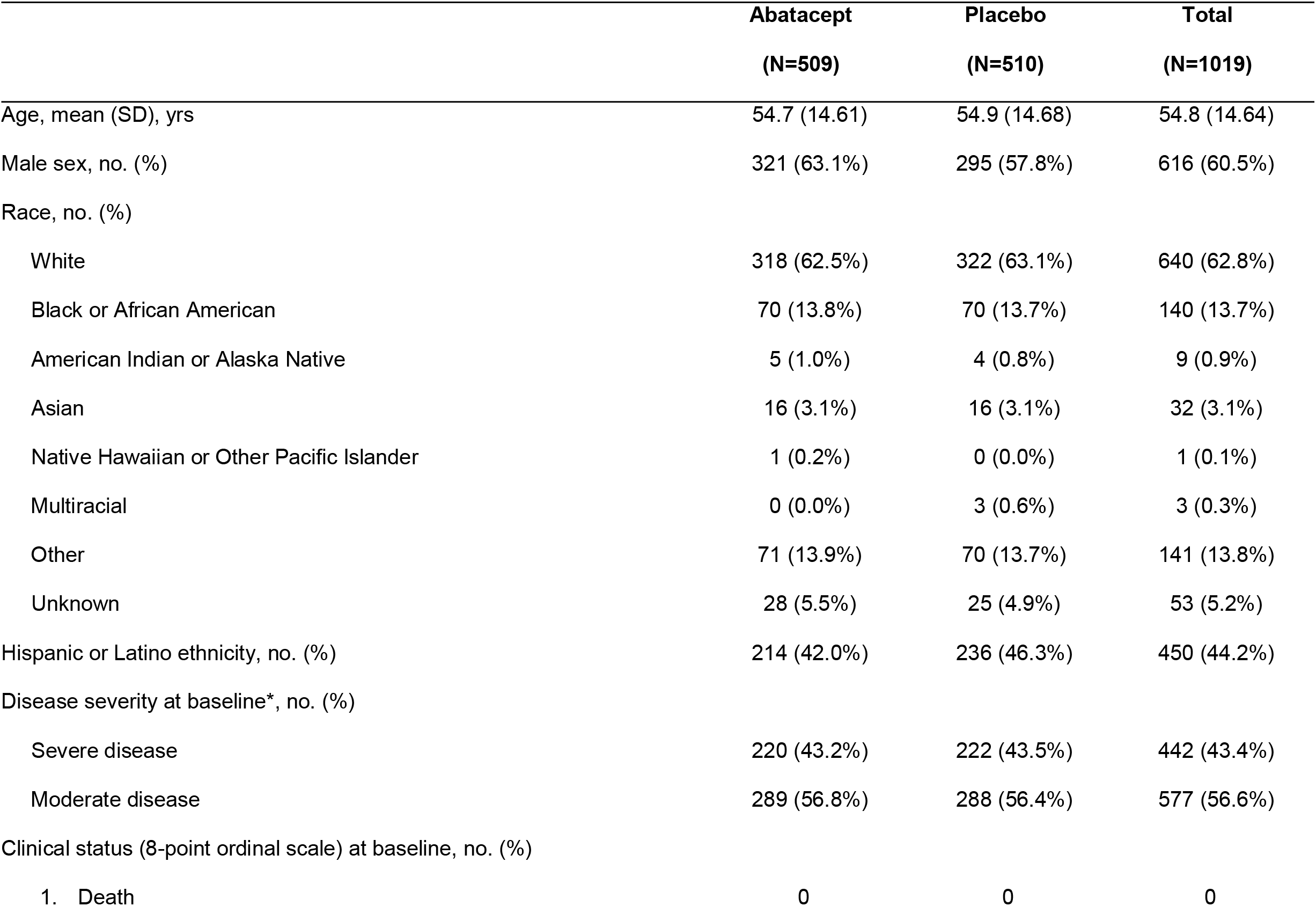
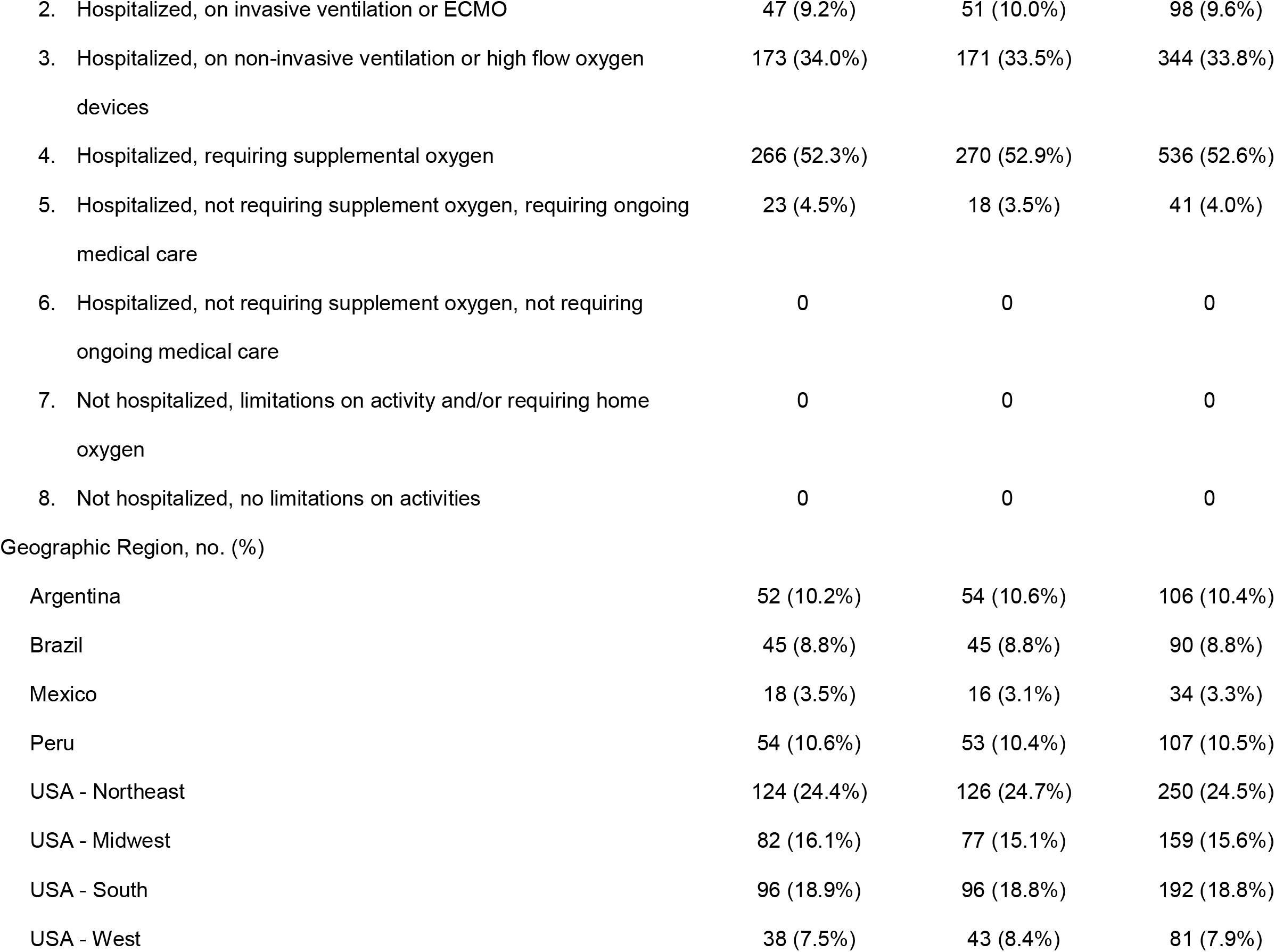
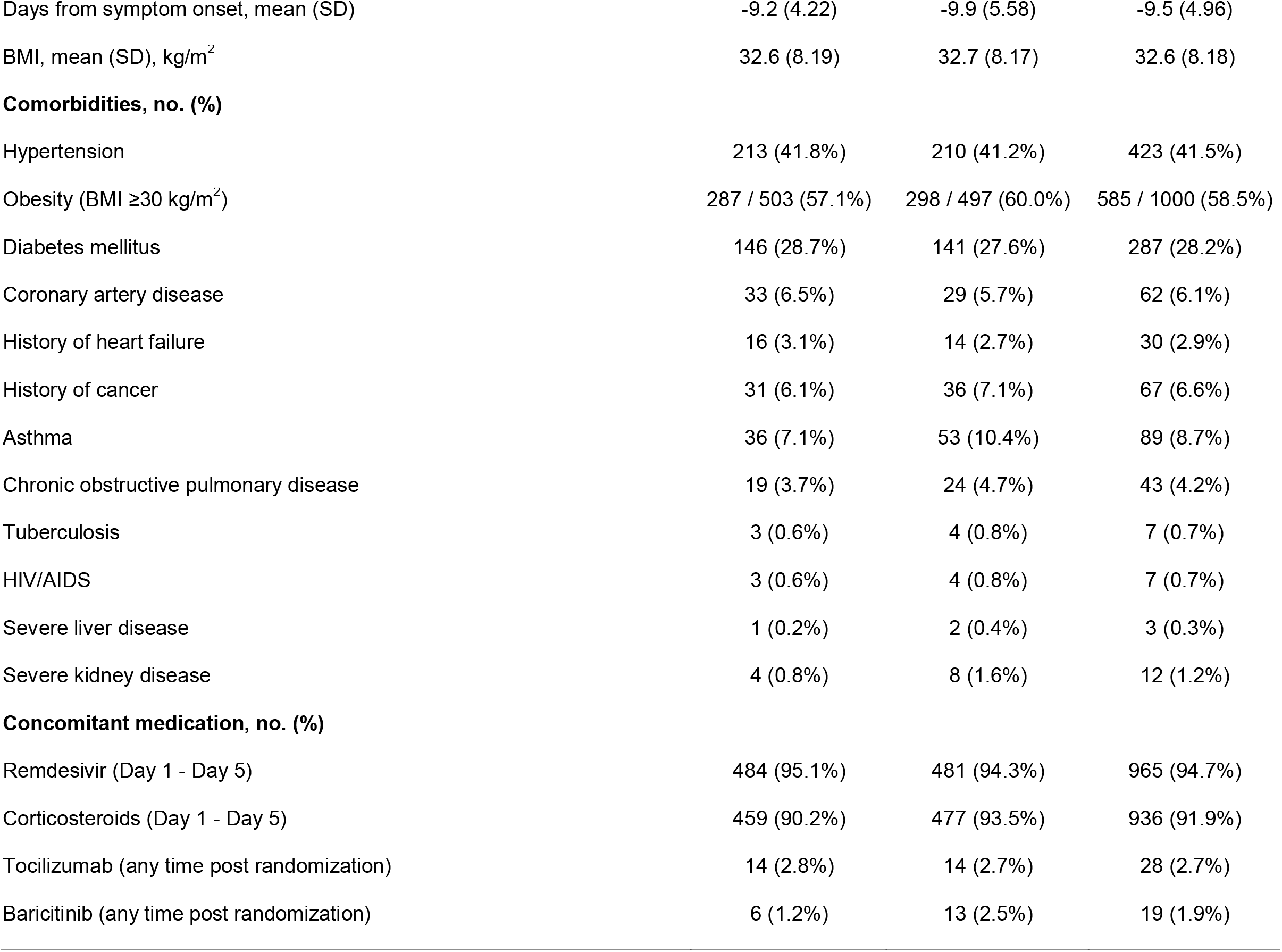
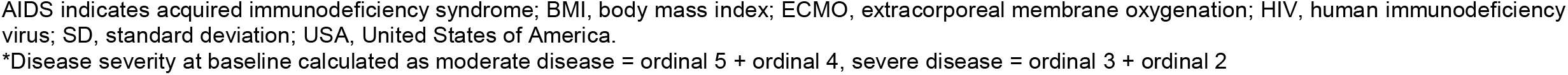
Demographics and baseline characteristics (modified intent-to-treat)

### Statistical Analysis

Sample size requirements were based on the ability to detect moderate improvement in time to recovery. A total of 788 recoveries were required to provide approximately 85% power to detect a recovery-rate-ratio (RRR) of 1.25 for abatacept versus placebo, assuming 73% of participants achieved recovery within 28 days.^16^ Primary efficacy analysis for abatacept versus placebo was based on the Fine-Gray model.^17^ The statistical analysis plan describes protection for Type 1 error for the primary endpoint and key secondary endpoints (28-day mortality and day 14 clinical status) using multiple testing procedures. Relevant p-value cutoffs are in **Table S1**. All-cause mortality through day 28 was analyzed as a binary endpoint with an indicator variable for treatment group through logistic regression. Clinical status through day 14 was calculated using a proportional odds model of improvement on an 8-point OS by ordinal logistic regression.

Participants who met eligibility were randomized in a two-stage process. Firstly, each participant was assigned with equal probability to one of the immunomodulatory agents (substudy) using an open-label design. Secondly, each participant was assigned in a blinded fashion to the test agent or its matching placebo in an n:1 ratio, where n equals the number of agents for which the participant was eligible. Participants in the placebo group were shared for each substudy once agent-specific eligibility criteria were applied. Thus, the shared placebo group utilized for the abatacept analysis was limited to participants who met enrollment criteria for the abatacept substudy.

### Safety Assessments

Safety assessments included a composite endpoint of death, serious adverse events (SAEs), or grade 3 (severe) and 4 (potentially life-threatening) adverse events (AEs) occurring through day 60. Secondary infections as adverse events of special interest through day 60 and discontinuation/suspension of trial-product administration were included.

## RESULTS

### Participants

A total of 1133 participants were eligible to receive abatacept; 524 were randomized to abatacept and 525 to shared placebo (**Figure S1**). Of those randomized, 509 (97.1%) received treatment as assigned in the abatacept group, and 510 (97.1%) in the shared placebo group, comprising the modified intention-to-treat (mITT) cohort. Participants were 60.5% male, mean (SD) age was 54.8 (14.6 [range 18, 98]) years, and mean body mass index was 32.6 (8.18) kg/m^2^. The cohort was 44.2% Hispanic or Latino, 62.8% White, and 13.7% Black (**Table 1**). There were 577 (56.6%) participants with moderate disease (no oxygen or low-flow oxygen) and 442 (43.4%) with severe disease (high-flow, noninvasive, or invasive ventilation/extracorporeal membrane oxygenation [ECMO]). Characteristics were similar among randomization groups (**Table 1, Table S2**).

Concomitant use of remdesivir and steroids was evenly distributed across randomized groups (**Table 1**). Use of EUA immunomodulators started after study treatment was low, with 28 (2.7%) receiving tocilizamab and 19 (1.9%) baricitinib post-randomization. Study treatment remained blinded.

### Primary Outcome

Participants in both groups had a median time-to-recovery of 9 days (IQR 8,10) with a RRR of 1.14 (95%CI 1.00–1.29; p=0.057), and a trend toward improvement with abatacept (**Figure S2**). Recovery rate estimates using a cumulative incidence function were 81.1% (95% CI 77.4–84.2) in the abatacept group and 76.8% (95% CI 72.9–80.3) in the shared placebo group. When analyzed according to severity of illness at randomization, the trend was strongest for participants on low-flow oxygen (OS 4) (RRR 1.14, 95% CI 0.97–1.35) and high-flow/noninvasive ventilation (OS 3) (RRR 1.24, 95% CI 0.96–1.59) (**Table 2, Figure S2**). Participants with C-reactive protein (CRP) >75 mg/L at randomization demonstrated the largest benefit, with a median of 10 days (IQR 9,12) to recovery with abatacept and 11 days (IQR 9, 15) with placebo (RRR 1.28, 95% CI 1.03–1.60), compared with participants with CRP ≤75 mg/L (interaction p=0.15) (**Table 2**). Those with a median duration of symptoms >9 days prior to randomization had a stronger benefit (RRR 1.36, 95% CI 1.10–1.68) than those earlier in the disease process. Participants with diabetes had a notable benefit from the use of abatacept (RRR 1.54, 95% CI 1.18–2.00) (**Figure S2**). No differences between geographic regions, or those receiving remdesivir or dexamethasone were observed.

**Table 2.**
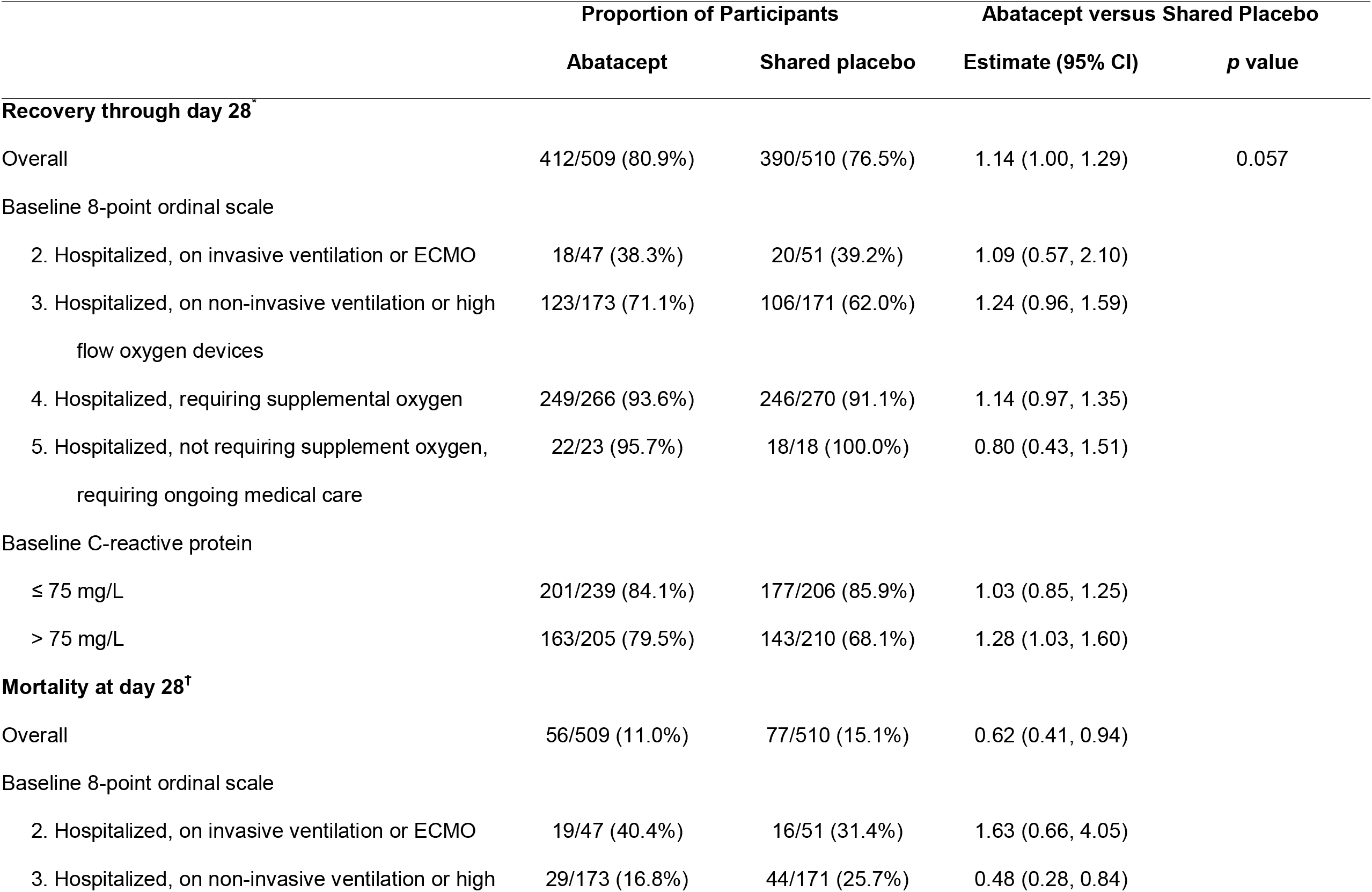
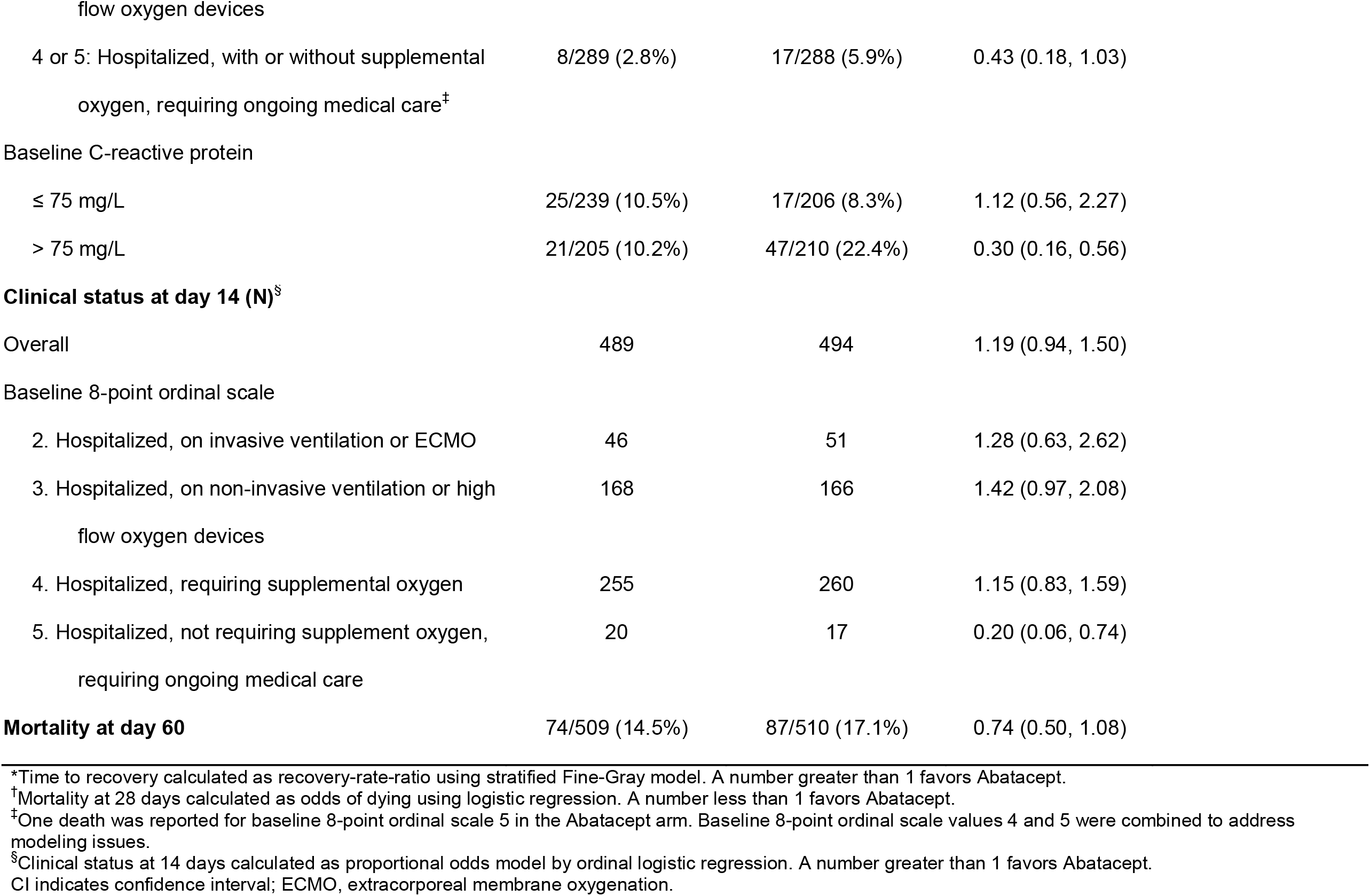
Primary and key secondary endpoints (modified intent-to-treat population)

|  | Proportion of Participants |  | Abatacept versus Shared Placebo |  |
| --- | --- | --- | --- | --- |
|  | Abatacept | Shared placebo | Estimate (95% CI) | *p* value |
| **Recovery through day 28<sup>\*</sup>** |  |  |  |  |
| Overall | 412/509 (80.9%) | 390/510 (76.5%) | 1.14 (1.00, 1.29) | 0.057 |
| Baseline 8-point ordinal scale |  |  |  |  |
| 2. Hospitalized, on invasive ventilation or ECMO | 18/47 (38.3%) | 20/51 (39.2%) | 1.09 (0.57, 2.10) |  |
| 3. Hospitalized, on non-invasive ventilation or high flow oxygen devices | 123/173 (71.1%) | 106/171 (62.0%) | 1.24 (0.96, 1.59) |  |
| 4. Hospitalized, requiring supplemental oxygen | 249/266 (93.6%) | 246/270 (91.1%) | 1.14 (0.97, 1.35) |  |
| 5. Hospitalized, not requiring supplement oxygen, requiring ongoing medical care | 22/23 (95.7%) | 18/18 (100.0%) | 0.80 (0.43, 1.51) |  |
| Baseline C-reactive protein |  |  |  |  |
| ≤ 75 mg/L | 201/239 (84.1%) | 177/206 (85.9%) | 1.03 (0.85, 1.25) |  |
| > 75 mg/L | 163/205 (79.5%) | 143/210 (68.1%) | 1.28 (1.03, 1.60) |  |
| **Mortality at day 28<sup>†</sup>** |  |  |  |  |
| Overall | 56/509 (11.0%) | 77/510 (15.1%) | 0.62 (0.41, 0.94) |  |
| Baseline 8-point ordinal scale |  |  |  |  |
| 2. Hospitalized, on invasive ventilation or ECMO | 19/47 (40.4%) | 16/51 (31.4%) | 1.63 (0.66, 4.05) |  |
| 3. Hospitalized, on non-invasive ventilation or high | 29/173 (16.8%) | 44/171 (25.7%) | 0.48 (0.28, 0.84) |  |

|  |  |  |  |
| --- | --- | --- | --- |
| flow oxygen devices |  |  |  |
| 4 or 5: Hospitalized, with or without supplemental oxygen, requiring ongoing medical care<sup>‡</sup> | 8/289 (2.8%) | 17/288 (5.9%) | 0.43 (0.18, 1.03) |
| Baseline C-reactive protein |  |  |  |
| ≤ 75 mg/L | 25/239 (10.5%) | 17/206 (8.3%) | 1.12 (0.56, 2.27) |
| > 75 mg/L | 21/205 (10.2%) | 47/210 (22.4%) | 0.30 (0.16, 0.56) |
| **Clinical status at day 14 (N)<sup>§</sup>** |  |  |  |
| Overall | 489 | 494 | 1.19 (0.94, 1.50) |
| Baseline 8-point ordinal scale |  |  |  |
| 2. Hospitalized, on invasive ventilation or ECMO | 46 | 51 | 1.28 (0.63, 2.62) |
| 3. Hospitalized, on non-invasive ventilation or high flow oxygen devices | 168 | 166 | 1.42 (0.97, 2.08) |
| 4. Hospitalized, requiring supplemental oxygen | 255 | 260 | 1.15 (0.83, 1.59) |
| 5. Hospitalized, not requiring supplement oxygen, requiring ongoing medical care | 20 | 17 | 0.20 (0.06, 0.74) |
| **Mortality at day 60** | **74/509 (14.5%)** | **87/510 (17.1%)** | **0.74 (0.50, 1.08)** |

\*Time to recovery calculated as recovery-rate-ratio using stratified Fine-Gray model. A number greater than 1 favors Abatacept.
<sup>†</sup>Mortality at 28 days calculated as odds of dying using logistic regression. A number less than 1 favors Abatacept.
<sup>‡</sup>One death was reported for baseline 8-point ordinal scale 5 in the Abatacept arm. Baseline 8-point ordinal scale values 4 and 5 were combined to address modeling issues.
<sup>§</sup>Clinical status at 14 days calculated as proportional odds model by ordinal logistic regression. A number greater than 1 favors Abatacept.
CI indicates confidence interval; ECMO, extracorporeal membrane oxygenation.

### Mortality

We observed 11.0% (56/509) 28-day all-cause mortality with abatacept compared with 15.1% (77/510) (odds ratio [OR] 0.62, 95% CI 0.41–0.94) with placebo, and 38% lower odds of death (**Figure 1, Figure 2A**). The overall trend for benefit of abatacept occurred across regions, and baseline characteristics (**Figure 1**). Subgroup analysis showed participants with moderate illness (OS 4/5) (OR 0.43, 95% CI 0.18–1.03) or high-flow/noninvasive ventilation (OS 3) (OR 0.48, 95% CI 0.28–0.84) had the strongest benefit of abatacept relative to placebo (**Figures 2C & D**). No benefit was found for participants enrolled on invasive ventilation/ECMO (OS 2) (OR 1.63, 95% CI 0.66–4.05) (**Figure 2E**). Participants not requiring oxygen at enrollment (OS 5) had a low event rate (1 death in the abatacept group). The interaction p-value across OS was p=0.06.

**Figure 1.**
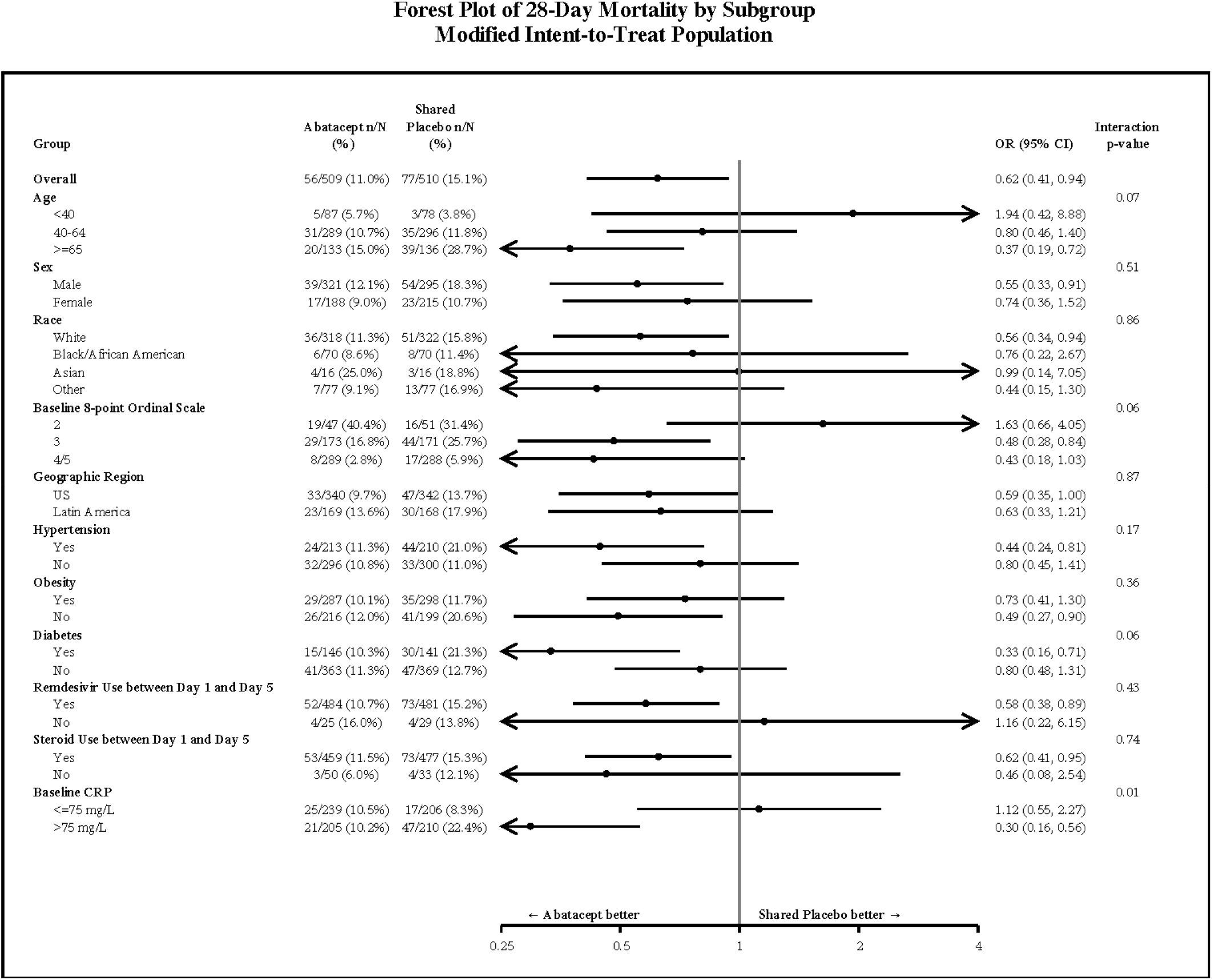
Forest plot of 28-day mortality by subgroup in the modified intent-to-treat population.

**Figure 2.**
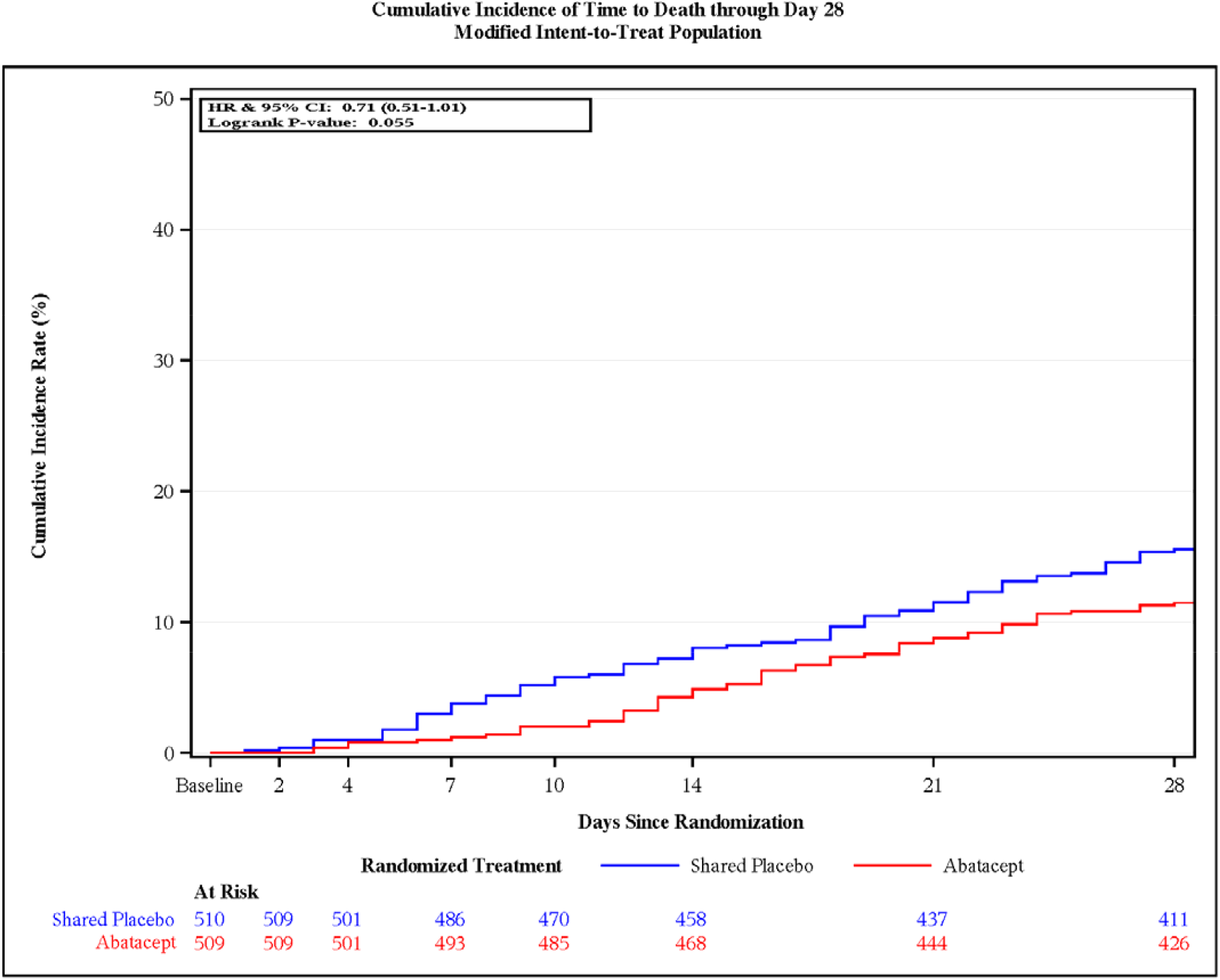
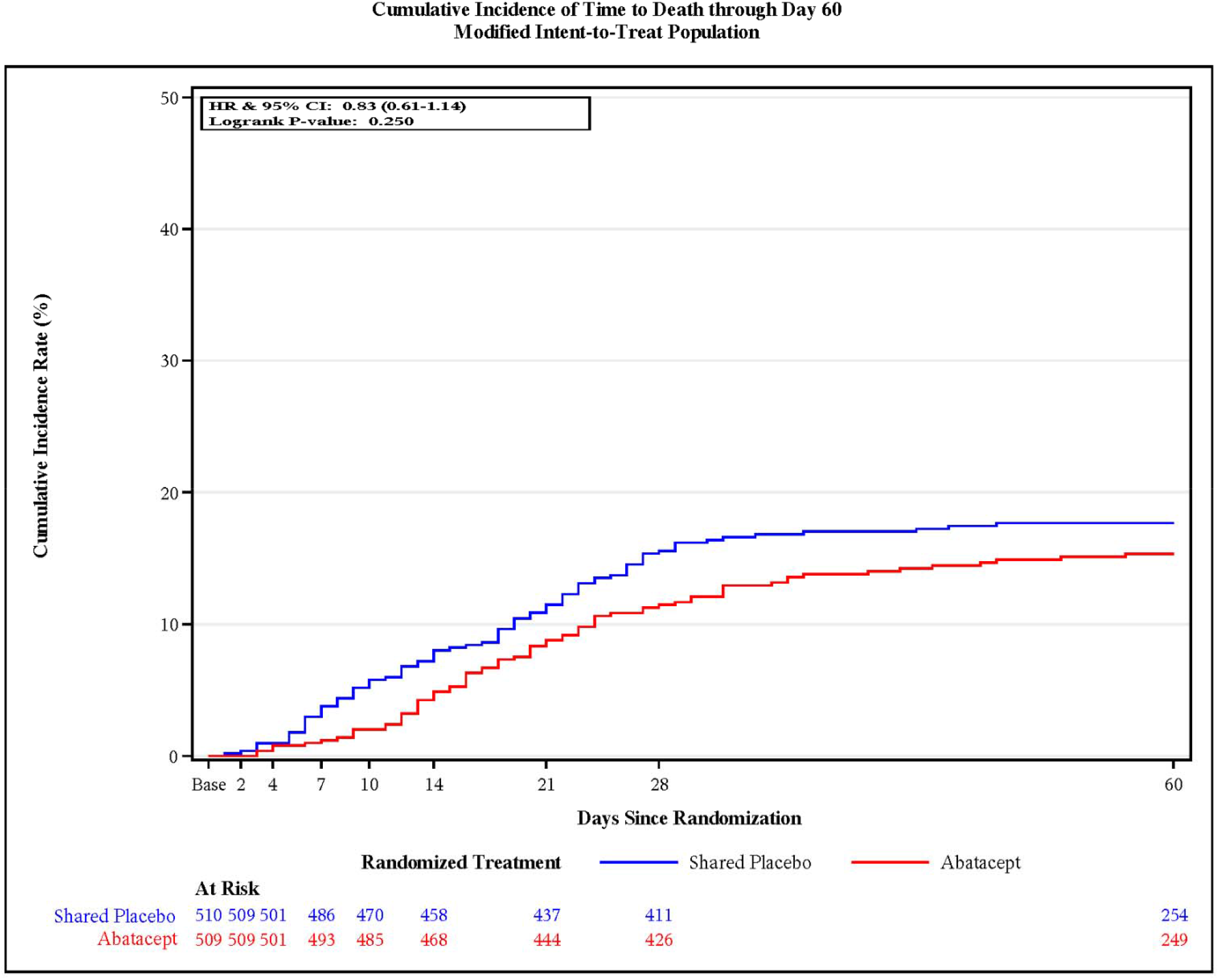
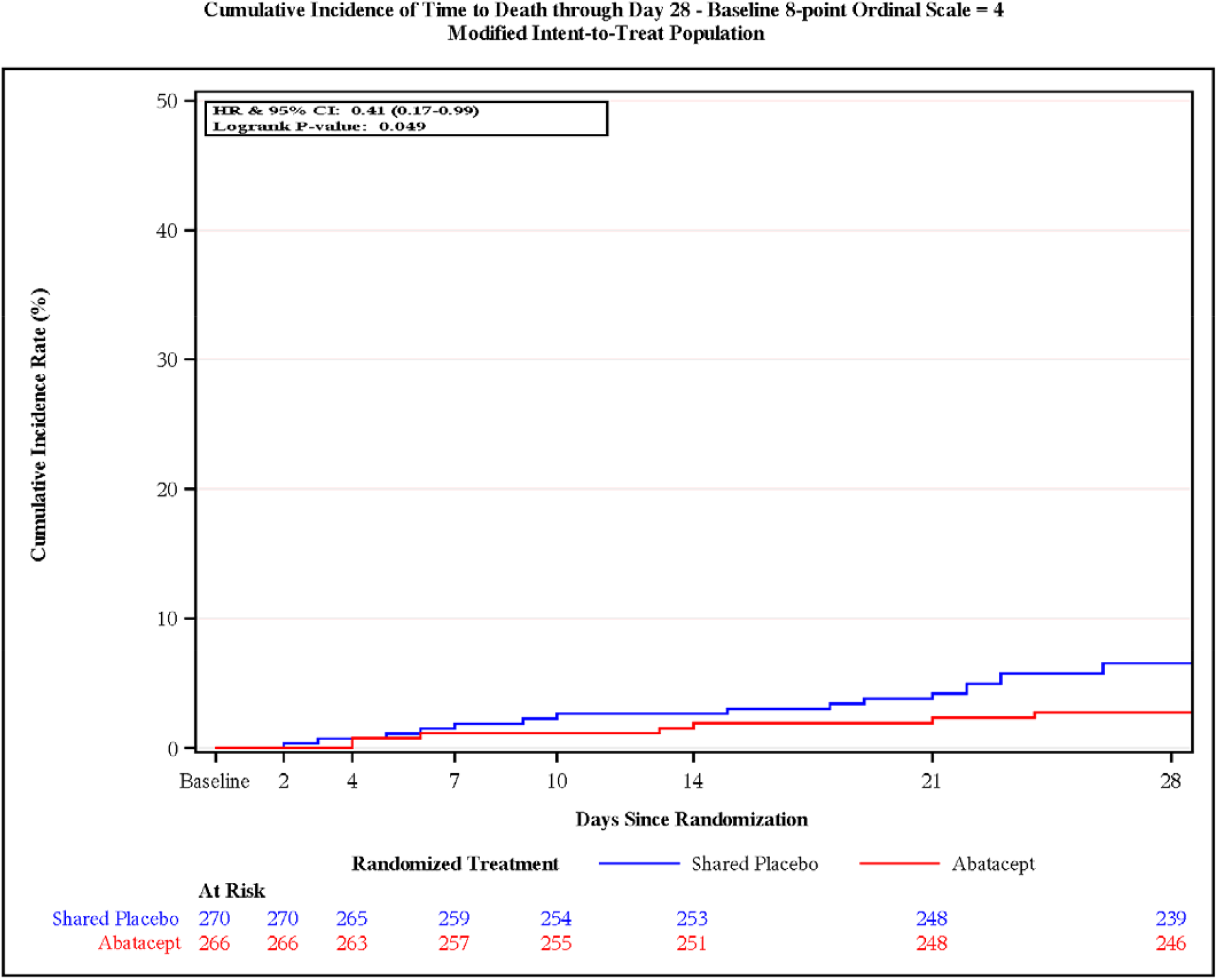
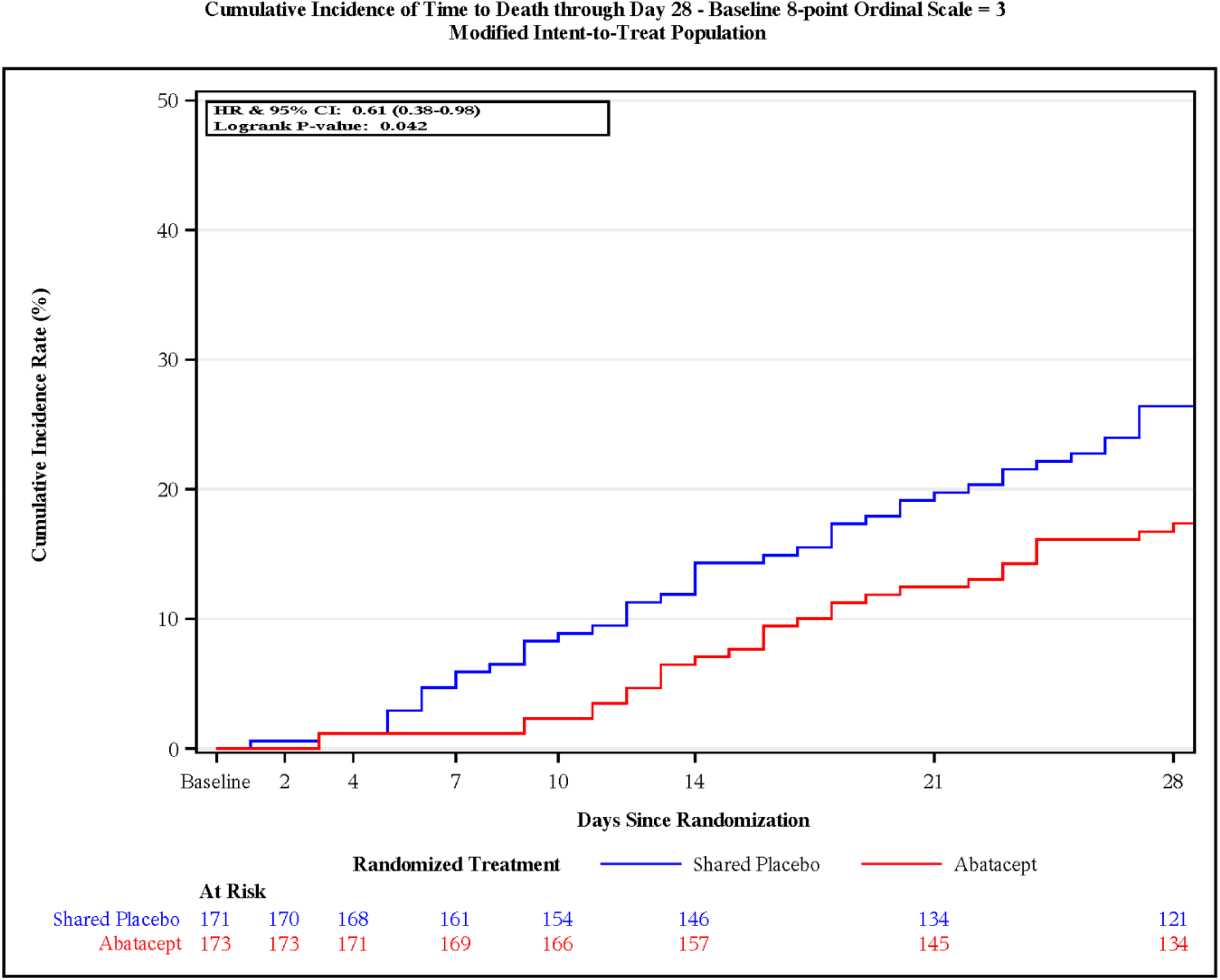
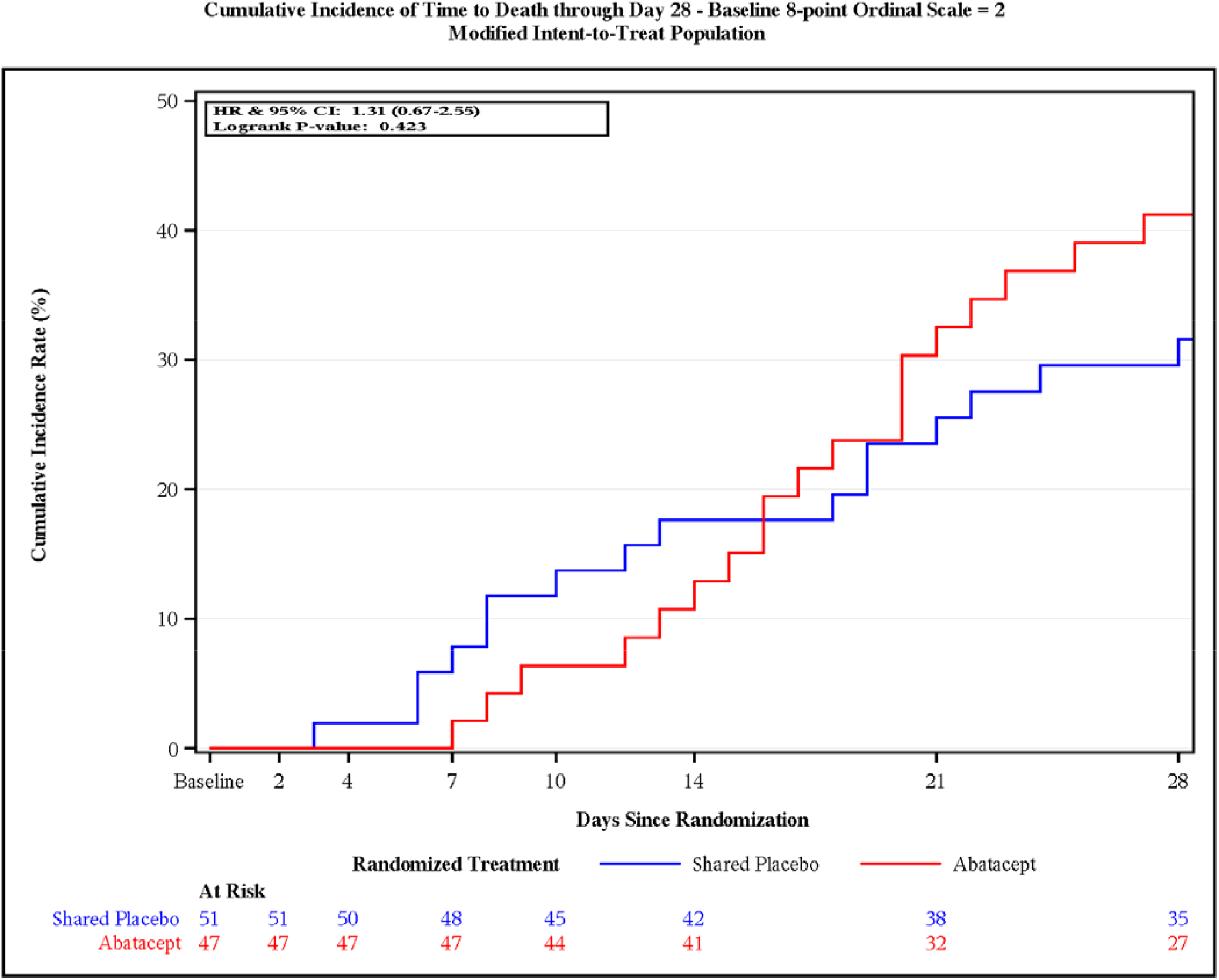
Kaplan-Meier curves demonstrating cumulative incidence of all-cause mortality (A) at 28 days, (B) at 60 days, (C) at 28 days for participants on low-flow oxygen (OS 4), (D) at 28 days for participants on high-flow oxygen or noninvasive ventilation (OS 3), and (E) at 28 days for participants mechanically ventilated or on ECMO (OS 2).

Participants with evidence of a systemic inflammatory response using a CRP cutoff of >75 mg/L demonstrated a stronger benefit (OR 0.30, 95% CI 0.16–0.56) compared with those with a CRP ≤75 mg/L (OR 1.12, 95% CI 0.55–2.27; interaction-p=0.006) (**Table 2**). As part of the 60-day safety assessment, we found 14.5% (74/509) mortality in the abatacept group and 17.1% (87/510) in the placebo group (OR 0.74, 95% CI 0.50–1.08) (**Table 2, Figure 2B**).

Twenty-eight participants died between day 29 and 60 (18 abatacept; 10 placebo). The majority (25/28) were intubated at day 28, although 3 had recovered and died after discharge.

### Clinical Status Change

We assessed proportional odds of improved clinical status at day 14 and 28 in the mITT population. The odds of improvement at day 14 were statistically unchanged with abatacept compared with placebo (OR 1.19, 95% CI 0.94–1.50) (**Figure S3**), but a substantial improvement was observed at day 28 (OR 1.35, 95% CI 1.06–1.73) (**Table S3**). All intention-to-treat data are presented in **Tables S4 and S5**.

### Safety Assessments

The composite safety endpoint at day 60 consisting of death, any SAE, and Grade 3/4 AEs indicated no difference for participants who received abatacept (33.2%) compared with placebo (34.9%) with a hazard ratio of 0.93 (95% CI 0.75–1.14; p=0.48) and a risk difference of -1.7% (**Table 3**). SAEs and Grade 3/4 AEs were not statistically different between the two groups. Any SAE was reported for 128 (25.1%) participants in the abatacept arm and 136 (26.7%) in the placebo arm. Nine (1.8%) participants in abatacept and 7 (1.4%) in the placebo group had an SAE related to study drug, none were deemed by the sponsor to be suspected unexpected serious adverse reaction related to abatacept administration. Three serious infusion reactions were reported in the abatacept arm and formally adjudicated. One grade 4 SAE met the case definition for anaphylaxis while two grade 3 SAEs met criteria for infusion reactions.^18^

**Table 3.**
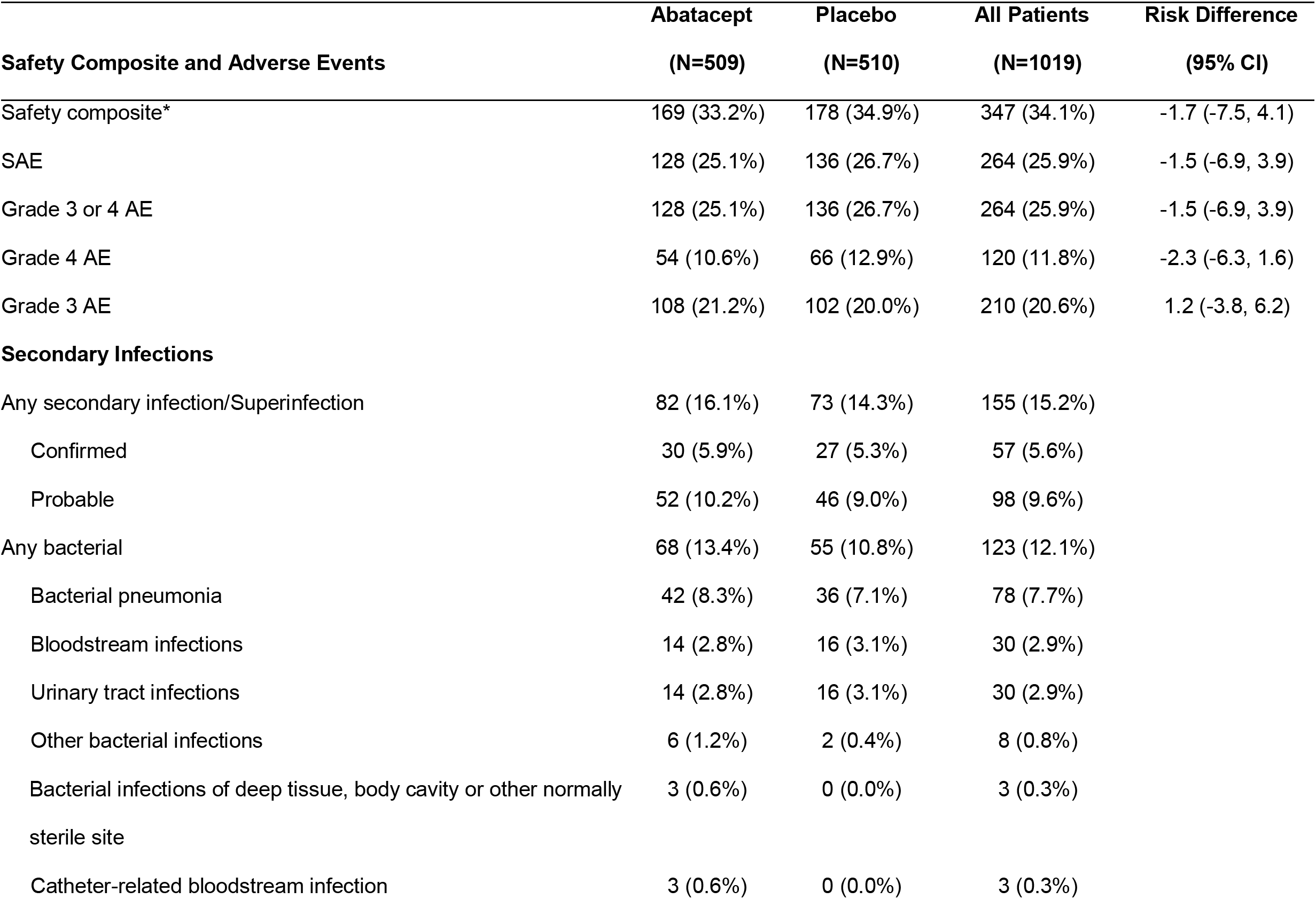
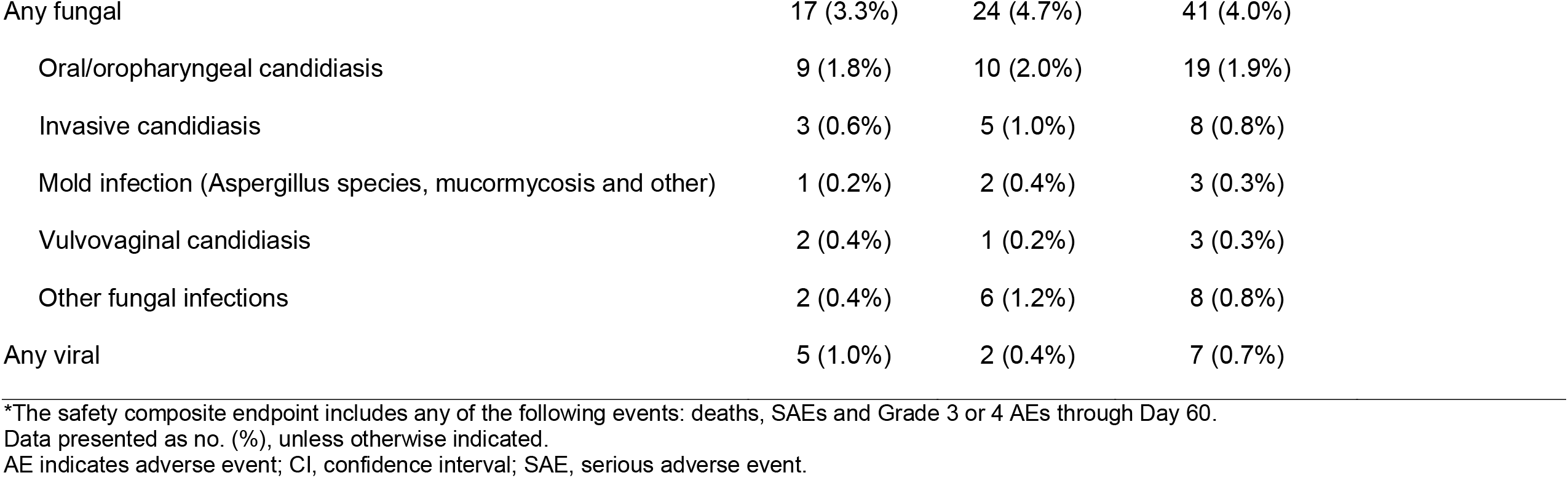
Safety composite and adverse events through day 60.

|  | **Abatacept** | **Placebo** | **All Patients** | **Risk Difference** |
| --- | --- | --- | --- | --- |
| **Safety Composite and Adverse Events** | **(N=509)** | **(N=510)** | **(N=1019)** | **(95% CI)** |
| Safety composite* | 169 (33.2%) | 178 (34.9%) | 347 (34.1%) | -1.7 (-7.5, 4.1) |
| SAE | 128 (25.1%) | 136 (26.7%) | 264 (25.9%) | -1.5 (-6.9, 3.9) |
| Grade 3 or 4 AE | 128 (25.1%) | 136 (26.7%) | 264 (25.9%) | -1.5 (-6.9, 3.9) |
| Grade 4 AE | 54 (10.6%) | 66 (12.9%) | 120 (11.8%) | -2.3 (-6.3, 1.6) |
| Grade 3 AE | 108 (21.2%) | 102 (20.0%) | 210 (20.6%) | 1.2 (-3.8, 6.2) |
| **Secondary Infections** |  |  |  |  |
| Any secondary infection/Superinfection | 82 (16.1%) | 73 (14.3%) | 155 (15.2%) |  |
| Confirmed | 30 (5.9%) | 27 (5.3%) | 57 (5.6%) |  |
| Probable | 52 (10.2%) | 46 (9.0%) | 98 (9.6%) |  |
| Any bacterial | 68 (13.4%) | 55 (10.8%) | 123 (12.1%) |  |
| Bacterial pneumonia | 42 (8.3%) | 36 (7.1%) | 78 (7.7%) |  |
| Bloodstream infections | 14 (2.8%) | 16 (3.1%) | 30 (2.9%) |  |
| Urinary tract infections | 14 (2.8%) | 16 (3.1%) | 30 (2.9%) |  |
| Other bacterial infections | 6 (1.2%) | 2 (0.4%) | 8 (0.8%) |  |
| Bacterial infections of deep tissue, body cavity or other normally sterile site | 3 (0.6%) | 0 (0.0%) | 3 (0.3%) |  |
| Catheter-related bloodstream infection | 3 (0.6%) | 0 (0.0%) | 3 (0.3%) |  |

|  |  |  |  |
| --- | --- | --- | --- |
| Any fungal | 17 (3.3%) | 24 (4.7%) | 41 (4.0%) |
| Oral/oropharyngeal candidiasis | 9 (1.8%) | 10 (2.0%) | 19 (1.9%) |
| Invasive candidiasis | 3 (0.6%) | 5 (1.0%) | 8 (0.8%) |
| Mold infection (Aspergillus species, mucormycosis and other) | 1 (0.2%) | 2 (0.4%) | 3 (0.3%) |
| Vulvovaginal candidiasis | 2 (0.4%) | 1 (0.2%) | 3 (0.3%) |
| Other fungal infections | 2 (0.4%) | 6 (1.2%) | 8 (0.8%) |
| Any viral | 5 (1.0%) | 2 (0.4%) | 7 (0.7%) |

\*The safety composite endpoint includes any of the following events: deaths, SAEs and Grade 3 or 4 AEs through Day 60. Data presented as no. (%), unless otherwise indicated. AE indicates adverse event; CI, confidence interval; SAE, serious adverse event.

Secondary infections by day 60 were reported for 82 (16.1%) and 73 (14.3%) participants in the abatacept and placebo arms. Notably, only a small number were confirmed by culture (30 [5.9%] abatacept vs. 27 [5.3%] placebo). Bacterial infection represented the majority of infections, with a small number of fungal or viral infections balanced between treatment arms (**Table 3**). Invasive fungal infections were rare, and occurred more often in the placebo group.

## DISCUSSION

This randomized, double-blind, placebo-controlled substudy evaluated use of single-dose intravenous abatacept in addition to standard of care therapy compared with placebo plus standard of care for the treatment of patients hospitalized with Covid-19. The trial substudy did not show a statistically significant benefit for addition of abatacept to standard of care for the primary endpoint of time-to-recovery. However, we found a substantial 28-day all-cause mortality benefit for abatacept, demonstrating 38% reduced odds of death for participants with moderate and severe disease. The mortality benefit was observed across different demographic groups, comorbidities, and independently from other Covid-19 therapies. Day 60 safety assessment showed preservation of the mortality benefit, although we did observe additional deaths in both abatacept and placebo groups. Subgroup analysis revealed that participants on oxygen or noninvasive ventilation and those with evidence of systemic inflammation (CRP >75 mg/L) realized a greater benefit from addition of a second immunomodulatory agent.

The nexus of overactive immune responses that are hallmarks of severe Covid-19 catalyzed researchers to identify effective anti-inflammatory agents to treat unregulated immune cascades early in the Covid-19 pandemic.^2,19^ Addition of dexamethasone was found to improve survival in patients requiring oxygen and became standard of care.^2^ However, the extraordinary morbidity and mortality of Covid-19 suggested the need for additional immunomodulators. The interleukin-6 (IL-6) antibody, tocilizumab, and the JAK1/JAK2 inhibitor, baricitinib, showed benefit for patients particularly in the setting of progressive respiratory failure.^20-22^ Results from these trials prompted the NIH to include tocilizumab or baricitinib as a second immunomodulator in addition to dexamethasone for patients with progressive respiratory failure and evidence of systemic inflammation (CRP>75 mg/L) in treatment guidelines.

The ACTIV-1 IM trial evaluated whether the use of additional immunomodulation by novel mechanisms of action improved outcomes for patients hospitalized with Covid-19, enrolling patients prior to successful trials on other secondary immunomodulators. We demonstrated a substantial mortality benefit for abatacept as well as infliximab in a parallel substudy. These data support the body of evidence for the use of a secondary immunomodulator in addition to dexamethasone, offer reassuring evidence for safety, and expand the armament of treatment options globally. Interestingly, the mortality benefit occurred across a wide spectrum of modes of action (selective T-cell costimulatory modulation, TNF-alpha inhibition, IL-6 inhibition, and JAK-1/2 inhibition), warranting further exploration of the role of inflammation in the pathogenesis of SARS-CoV-2.

Important knowledge gaps remain, particularly in the setting of patients requiring conventional oxygen supplementation when first hospitalized. Subgroup analyses from the ACTIV-1 IM trial begin to address these critical questions. We show a mortality benefit in both moderate and severe disease across two distinct immunomodulatory agents (abatacept reported here and infliximab reported separately). Our results for patients with moderate illness show improvement independent of inflammatory markers or clinical factors. Findings for moderate illness observed across two agents studied in ACTIV-1 IM suggest treatment with abatacept, and other immunomodulators, early in the disease process could provide benefit.

We demonstrate safety and benefit of additional immunomodulation in conjunction with dexamethasone for patients on conventional oxygen with Covid-19, but patients may have heterogenous inflammatory and treatment responses. Clinicians balance risk over benefit of additional immunomodulators for individual patients. Our subgroup analysis showed an enrichment of both time to recovery and mortality benefit in participants with CRP >75 mg/L, suggesting biomarkers could identify participants at highest risk of deterioration. Heterogeneity of pathogenesis also exists for patients on invasive ventilation/ECMO, demonstrating both hyper- and hypo-inflammatory responses.^23-26^ The lack of benefit shown for abatacept in these participants could be influenced by the disease variability at this stage. Exploring markers of therapeutic response, both early and late in illness, will improve our treatment approach to hospitalized Covid-19 patients.**Error!**

### Bookmark not defined

Addition of abatacept to the collections of medications effective for treatment of Covid-19 represents a key step to expanded access and safety. It is a well-characterized and tolerated medication with few precautions related to its use. Abatacept targets the T-cell costimulatory signaling that fuels the hyperinflammatory response, a unique mechanism among immunomodulators that could bring harmony to the immune system while preserving viral clearance. Robust safety data monitored to day 60 ensured capture of all events, and did not demonstrate a statistically significant increase in infection or other SAEs.

The primary endpoint of time to recovery did not reach statistical significance. Based on use of the gatekeeping approach, key secondary endpoints cannot be considered statistically significant even with a nominal p-value <0.05 for 28-day mortality. Nonetheless, improved 28-day mortality represents a considerably important finding clinically. The emergence of SARS-CoV-2 created an unprecedented opportunity to improve our understanding of pathogenesis and therapeutic approaches to pandemic viral pathogens, albeit in the setting of rapidly evolving variants and clinical practices. Determination of the most appropriate primary endpoint in the midst of a pandemic has been a challenge for many Covid-19 trials.^27^ As the field progresses, alternative primary endpoints will likely incorporate accumulated knowledge to improve assessment of clinically relevant endpoints.^27,28^

As the landscape of immunity and SARS-CoV-2 variants changes, an understanding of best treatment practices for hospitalized patients remains a priority. Therapeutic approaches to manage patients hospitalized with Covid-19 will likely require agents with several different mechanisms of action. Improved approaches to identifying patients most likely to benefit from therapies, especially at the bookends of early and late disease, could be advantageous as the pandemic evolves. This report from the ACTIV-1 IM master protocol shows that the addition of abatacept to standard of care provides a substantial mortality benefit and minimal additional risk for patients requiring oxygen or noninvasive ventilation.

## Data Availability

It is expected that a partial de-identified patient dataset will be made available via the ACTT Trial Database at NIAID (Access Clinical Data @ NIAID) beginning 6 months after publication of the primary results and for 5 years following article publication.

## Acknowledgments

We thank the members of the ACTIV-1 Study Team (see the Supplementary Appendix) for their many contributions in conducting the trial, the members of the data and safety monitoring board, William C. Blackwelder, PhD (chair) (University of Maryland School of Medicine, Baltimore, MD), Timothy G. Buchman, PhD, MD (Emory University School of Medicine, Atlanta, GA), Wilbur H. Chen, MD MS (University of Maryland School of Medicine, Center for Vaccine Development and Global Health, Baltimore, MD), Lawrence H. Moulton PhD (Johns Hopkins University, Bloomberg School of Public Health, Baltimore, MD), David M. Parenti, MD (The George Washington University School of Medicine, Washington, DC), Carol O. Tacket, MD (University of Maryland School of Medicine, Center for Vaccine Development and Global Health, Baltimore, MD), William Checkley, MD, PhD Johns Hopkins University, Baltimore, MD), Beatriz Grinsztejn, MD (Instituto Nacional de Infectologia Evandro Chagas-Fiocruz, Rio de Janeiro, Brazil) for their oversight; the Community Advisory Board, Larisa Caicedo, Ashish Cowlagi, Anna Davis, Lincoln Larmond, Doug Lindsey, Bob Pearson and the patients and their families for their altruism in participating in this trial.

## Funding

This project has been funded with federal funds from the U.S. Department of Health and Human Services, Office of the Assistant Secretary for Preparedness and Response, Biomedical Advanced Research and Development Authority, under contract number HHSO100201400002I/75A50120F33002. The findings and conclusions in this publication are those of the authors and do not necessarily represent the views of the Department of Health and Human Services or its components. Bristol Myers Squibb provided abatacept for use in this trial but did not provide any financial support. Gilead Sciences provided remdesivir for use in this trial but did not provide any financial support. Employees of BMS and Gilead Sciences participated in discussions about protocol development and in weekly protocol team calls. The final trial protocol was developed by the protocol chair, Dr. William Powderly, the IND sponsor Dr. Daniel K. Benjamin, Jr, and a protocol development committee including representatives from the National Center for Advancing Translational Sciences (NCATS). NCATS had a collaborative role in the trial design, management, interpretation of the data and the preparation of the manuscript along with the protocol chair, the study statisticians and the study writing group.

Infrastructure support and resources for research reported in this publication were provided in part by NCATS of the National Institutes of Health under award number(s):UL1TR002345 to Washington University in St. Louis; the Duke University – Vanderbilt University Medical Center Trial Innovation Center (U24TR001608) (NCATS, Trial Innovation Network).

## Data Sharing

A data sharing statement provided by the authors is available with the full text of this article.

## Notes

### Competing Interest Statement

This information is being collected.

### Clinical Trial

NCT04593940

### Author Declarations

All sites conducted the study under IRB/EC oversight. WCG IRB gave ethical approval for this work in the US for all sites with the exception of one site that received approval from the Mid-Michigan IRB of the MyMichigan Medical Center. The following ethics committees gave approval in Argentina-Fundacion Medica del Litoral (FUMELIT), Comite de etica en investigacion del sanatorio Britanico, CEDIMP-Comite de Etica Instituto Medico Platense, Comite de Etica en Investigacion del Hospital Gral de Agudos J. M. Ramos Mejia, Comite de etica e investigacion clinica (CEIC), Sanatorio Allende, Comite de Etica en Investigacion (CIEIS) del Nino y del Adulto, Comite de Etica en Investigacion con Seres Humanos del Hospital Interzonal General Dr. Jose Penna. The following ethics committees gave approval in Brazil-Comite de Etica em Pesquisa da Irmandade da Santa Casa de Misericordia de Porto Alegre, Comite de Etica da Associacao dos Funcionarios Publicos do RS-Hospital Ernesto Dornelles, Comite de Etica em Pesquisa do Hospital de Clinicas de Porto Alegre, Comite de Etica em Pesquisa em Seres Humanos da Pontificia Universidade Catolica de Campinas PUC Campinas, Comite de Etica em Pesquisa do Instituto DOr de Pesquisa e Ensino-IDOR, Comite de Etica em Pesquisa do Hospital Felicio Rocho, Comite de Etica em Pesquisa do Hospital 9 de Julho. The following ethics committees gave approval in Mexico-Nuevo Hospital Civil de Guadalajara Dr. Juan I. Menchaca, Hospital Universitario Dr. Jose Eleuterio Gonzalez, Centro Medico Nacional Siglo XXI (Coordinacion de Investigacion en Salud), Hospital Civil de Guadalajara Fray Antonio Alcalde. The following ethic committees gave approval for all sites in Peru-Comite Nacional Transitorio de Etica e Investigacion (CNTEI).

