## Supplementary Appendix for "Abatacept for Treatment of Adults Hospitalized with Moderate or Severe Covid-19"

Additional members of the study group includes:

**United States Sites**

Duke University Medical Center, Durham, NC. Cameron Wolfe, M.B.B.S., M.P.H.

Jamaica Hospital Medical Center, Jamaica, NY. Mahendra Patel, M.D.

Virginia Commonwealth University Medical Center, Richmond, VA. Arun Sanyal, M.D.

University of Tennessee Medical Center, Knoxville, TN. Jason Green, D.O.

Providence Medical Research Center, Spokane, WA. Radica Alicic, M.Sc., M.D.

University of Oklahoma Health Sciences Center, Oklahoma City, OK. Huimin Wu, M.D., M.P.H.,

Boston Medical Center, Boston, MA. Benjamin Linas, M.D., M.P.H.

Stanford University Medical Center, Palo Alto, CA. Philip Grant, M.D.

Mayo Clinic, Rochester, MN. Vivek Iyer, M.D.

UCLA Medical Center, Los Angeles, CA. Otto Yang, M.D.

Hackensack University Medical Center, Hackensack, NJ. Bindu Balani, M.D.

New York University Langone Medical Center, New York, NY. Sam Parnia, M.D.

University of Arkansas Medical Sciences, Little Rock, AR. Ryan Dare, M.D., M.S.

Wake Forest University Health Sciences, Winston-Salem, NC. Caryn G. Morse, M.D., M.P.H.

University of Utah, Salt Lake City, UT. Estelle S. Harris, M.D.

MedStar Washington Hospital Center, Washington, DC. Glenn Wortmann, M.D.

Tufts Medical Center, Boston, MA. Nicholas Hill, M.D.

UF Health Research, Jacksonville, FL. Shama Patel, M.D.

Ochsner Medical Center, New Orleans, LA. Julia Garcia-Diaz, M.D.

Riverside University Health System, Moreno Valley, CA. Suman Thapamager, M.D.

University of Texas Health Science Center- Tyler, Tyler, TX. Megan Devine, M.D.

Tulane University Health Science Center, New Orleans, LA. Christine M. Bojanowski, M.D., M.S.C.R.

Anne Arundel Medical Center, Annapolis, MD. Barry Meisenberg, M.D.

University of Mississippi Medical Center, Jackson, MS. Gailen Marshall, M.D.

University of Missouri Health System, Columbia, MO. Dima Dandachi, M.D.

Gundersen Health System, La Crosse, WI. Arick Sabin, D.O.

Avera McKennan Hospital & University Health Center, Sioux Falls, SD. Anthony Breemo, M.D.

Trinity Mother Frances Hospital, Tyler TX. Suman Sinha, M.D.

University of Washington Medical Center, Seattle, WA. Christopher Goss, M.D.

West Virginia University, Morgantown, WV. Rebecca Reece, M.D.

Mercy St. Vincent Medical Center, Toledo, OH. Arlette Aouad, M.D.

University of Buffalo, Buffalo, NY. Seth Glassman, M.D.

University of Kentucky, Lexington, KY. Peter Morris, M.D.

The University of Texas Health Science Center – Houston, Houston, TX. Bela Patel, M.D.

University of Texas - Rio Grande Valley, Weslaco, TX. Fatimah Bello, M.D.

**Ex-US Sites**

Hospital Ernesto Dornelles, Porto Alegre, Brazil. Juliana Cardozo Fernandes,M.D., M.Sc.

Hospital de Chancay, Lima, Peru. Oscar Carbajal, M.D.

Hospital Rawson, Cordoba, Argentina. Lorena Ravera M.D.

Hospital Felcio Rocho, Belo Horizonte, Brazil. Mozar Castro M.D.

Hospital Regional Lambayaque, Chiclayo, Peru. Miguel Villegas-Chiroque, M.D.

Sanatorio Allende, Cordoba, Argentina. Fernando Oscar Riera, M.D.

Hospital Universitario Dr. Jose Eleuterio Gonzalez, Nuevo Leon, Mexico. Adrian Camacho, M.D.

Santa Casa de Misericordia de Porto Alegre, Porto Alegre, Brazil. Claudio Stadnik M.D.

Hospital Nacional Arzobispo Loayza, Lima, Peru. Jorge Gave, M.D.

Hospital Brasilia, Brasilia, Brazil. Rodrigo Biondi. M.D.

Hospital Santa Rosa, Piura, Peru. Ronal Gamarra Velarde, M.D.

Instituto D'OR de Ensino e Pesquisa, Rio de Janeiro, Brazil. Jose Cerbino Neto, M.D.

Hospital Interzonal Dr Jose Penna Bahia Blanca, Bahia Blanca, Argentina. Juan Ditondo, M.D.

Hospital Ramos Mejia, Buenos Aires, Argentina. Marcelo H. Losso, M.D.

**Other Contributors**

AbbVie, North Chicago, IL. William Ferguson, PhD

Bristol Myers-Squibb, New York, NY. Jonathan Sadeh, MD MSc

Janssen Pharmaceuticals/Johnson and Johnson, Beerse, Belgium. Richard Melsheimer

Gilead, Foster City, CA. Huyen Cao, MD

Duke Clinical Research Institute (DCRI), Durham, NC. Rose Beci, Adam Silverstein, Sean O’Brien, PhD

Technical Resources International, Inc (TRI), Bethesda, MD. Daniel Molina, PharmD, MBA; Sandhya Rao, MD, M.Ch

Health and Human Services, Washington, DC. Thomas Stock, William Erhardt, M.D. (H-CORE, formally Operation Warp Speed), Sarah Read,

National Center for Advancing Translational Sciences, Bethesda, MD. Jessica Springer, RN, MPH/MPA,

### **ACTIV-1 Study Team Members**

The ACTIV-1 study was a significant collaborative effort across many countries, partners and sites. Below is a list of sites and staff that worked tirelessly to implement and execute the ACTIV-1 Trial.

**United States Sites**

Washington University St. Louis, St Louis, MO. William G. Powderly, M.D., Jane A. O’Halloran, M.D., Rachel Presti, M.D., Ph.D., Adriana Rauseo Acevedo, M.D., Luis Parra Rodriguez, M.D.

Duke University Health System, Durham, NC. Cameron R. Wolfe, M.B.B.S, M.P.H., Emily R. Ko, M.D., Ph.D., Tatyana Der, M.D., M.H.Sc., Lana Wahid, M.D., John J. Engemann, M.D., Gloria Pinero, Beth McLendon-Arvik, R.Ph., Lynn Whitt, R. Ph., Jenny Shroba, R. Ph.

Weill Cornell Medical College, New York, NY. Matthew W. McCarthy, M.D., Elizabeth Salsgiver, Kate Willsey, Candace Alleyne, Britta Witting.

Robert Wood Johnson Medical School, New Brunswick, NJ. Reynold Panettieri, M.D.

Jamaica Hospital Medical Center, Jamaica, NY. Mahendra Patel, M.D.

Oregon Health and Science University, Portland, OR. Akram Khan, M.D.

University of Illinois at Chicago, Chicago, IL. Alfredo J. Mena Lora, M.D., Rodrigo Burgos, Pharm.D., Brenna Lindsey, M.P.H., Fischer Herald, Pharm.D.

University of Kansas Medical Center, Kansas City, KS. Matthias Salathe, M.D., Charles D. Bengtson, M.D., Andreas Schmid, M.D., Kimberly Lovell, Ph.D.

St. Lawrence Health System, Potsdam, NY. Eyal Kedar, M.D., Carly Lovelett, C.C.R.P., M.S., M.B.A., Daniel Soule, D.O., Daniel Jaremczuk, C.C.R.P.

Trinitas Hospital, Elizabeth, NY. Gerardo Capo, M.D.

University of Texas Health Science Center at San Antonio, San Antonio, TX. Thomas F. Patterson, M.D., Heta Javeri, M.D., Philip O. Ponce, M.D., Danielle O. Dixon, D.O.

University of Rochester Medical Center-Strong Memorial Hospital, Rochester, NY. Christopher Palma, M.D., Sc.M., Caroline M. Quill, M.D.

University of North Carolina Hospital, Chapel Hill, NC. Anne Lachiewicz, M.D.

Virginia Commonwealth University Medical Center, Richmond, VA. Arun Sanyal, M.D.

MidMichigan Medical Center – Midland, Midland, MI. John Blamoun, M.D.

University of Tennessee Medical Center, Knoxville, TN. Jason Green, D.O., Abby Smith, R.N., Glenda Brown, R.N.

Providence Medical Research Center, Spokane, WA. Radica Alicic, M.Sc., M.D., Kyle Varner, M.D., Joni Baxter, R.N., Tracy Roundy, B.S.

University of Massachusetts Medical Center, Worcester, MA. Jennifer P. Wang, M.D., Mary Co, M.D., Mireya Wessolossky, M.D., Juan Perez-Velazquez, M.D.

University of Oklahoma Health Sciences Center, Oklahoma City, OK. Huimin Wu, M.D., M.P.H., Jennifer Holter-Chakrabarty, M.D., Brittany Karfonta, Juvaria Anjum.

Boston Medical Center, Boston, MA. Benjamin Linas, M.D., M.P.H., Jai Marathe, M.D., Myriam Castagne, Daniel MomPoint.

Stanford University Medical Center, Palo Alto, CA. Philip Grant, M.D.

Mayo Clinic, Rochester, MN. Vivek Iyer, M.D.

UCLA Medical Center, Los Angeles, CA. Otto Yang, M.D.

Hackensack University Medical Center, Hackensack, NJ. Bindu Balani, M.D., Marylynn Breslin.

New York University Langone Medical Center, New York, NY. Sam Parnia, M.D.

University of Arkansas Medical Sciences, Little Rock, AR. Ryan Dare, M.D., M.S., Susan Dodson, M.B.A., B.S.N., R.N.

Wake Forest University Health Sciences, Winston-Salem, NC. Caryn G. Morse, M.D., M.P.H., John Williamson, Pharm.D.

University of Utah, Salt Lake City, UT. Estelle S. Harris, M.D., Elizabeth A. Middleton, M.D. MedStar Washington Hospital Center, Washington, DC. Glenn Wortmann, M.D.

Tufts Medical Center, Boston, MA. Nicholas Hill, M.D.

UF Health Research, Jacksonville, FL. Shama Patel, M.D.

Ochsner Medical Center, New Orleans, LA. Julia Garcia-Diaz, M.D.

Riverside University Health System, Moreno Valley, CA. Suman Thapamager, M.D.

University of Texas Health Science Center- Tyler, Tyler, TX. Megan Devine, M.D.

Tulane University Health Science Center, New Orleans, LA. Christine M. Bojanowski, M.D., M.S.C.R.

Anne Arundel Medical Center, Annapolis, MD. Mai Tavadze, M.D., Barry Meisenberg, M.D.

University of Mississippi Medical Center, Jackson, MS. Gailen Marshall, M.D.

University of Missouri Health System, Columbia, MO. Dima Dandachi, M.D.

Gundersen Health System, La Crosse, WI. Arick Sabin, D.O.

Avera McKennan Hospital & University Health Center, Sioux Falls, SD. Anthony Breemo, M.D., Nate Miller, M.D.

Trinity Mother Frances Hospital, Tyler TX. Suman Sinha, M.D.

University of Washington Medical Center, Seattle, WA. Christopher Goss, M.D.

West Virginia University, Morgantown, WV. Rebecca Reece, M.D.

Mercy St. Vincent Medical Center, Toledo, OH. Arlette Aouad, M.D.

University of Buffalo, Buffalo, NY. Seth Glassman, M.D.

University of Kentucky, Lexington, KY. Peter Morris, M.D.

The University of Texas Health Science Center – Houston, Houston, TX. Bela Patel, M.D., Pratik Doshi, M.D.

University of Texas - Rio Grande Valley, Weslaco, TX. Fatimah Bello, M.D.

**Ex-US Sites**

Hospital Nacional Hipolito Unanue, Lima, Peru. Patricia Segura Nuñez,M.D., Pablo Torres, M.D., Ricardo Sanchez, M.D., Gladys Murga, M.D.

Hospital Central de la Fuerza Aerea del Peru, Lima, Peru. Sabina Mendiviltuchia de Tai, M.D., Johana Calderon Galvez, M.D., Maria Lyda Icochea Perez, M.D., Claudia Carolina Becerra Nunez, M.D.

Hospital Ernesto Dornelles, Porto Alegre, Brazil. Juliana Cardozo Fernandes,M.D., M.Sc., Felipe Ceriolli Breda, M.D., Mauricio Mello Roux Leite, M.Sc., Tobias Milbradt, M.Sc.

Sanatorio Diagnostico, Santa Fe, Argentina. Martin Maillo, M.D., Luz Rodeles, M.D., Ph.D., Nadia Benzaquen, M.D., Sebastian Pezzini, M.D.

Nuevo Hospital Civil de Guadalajara Juan I. Menchaca, Guadalajara, Mexico. Daniel Rodríguez Gonzalez, M.D., Iliana Higareda Almaraz, M.D., Víctor Hugo Madrigal Robles, M.D., María Fernanda Rosas Ismerio, M.D.

Clinica Central S.A., Villa Regina, Argentina. Horacio Ariza, M.D., Juan Manuel Calderon, M.D., Eduardo Borsetta, M.D., Noemí Sandoval, B.Q.

Hospital e Maternidade Celso Pierro - PUC Campinas, Campinas, Brazil. Maria Patelli Juliani Souza Lima, M.D.

Hospital de Clinicas de Porto Alegre HCPA, Porto Alegre, Brazil. Eduardo Sprinz, M.D., Laura De Bona,B.Q., Maria Eduarda Claus, Nut., Arthur Pille, M.D.

Sanatorio Británico, Rosario, Argentina. Esteban Nannini, M.D., Matías Lahitte, M.D., Mariángeles Fenés M.D., Cecilia Bianchi M.D., María Emilia Miserere M.D.

Instituto Medico Platense, La Plata, Argentina. Analia Mykietiuk, M.D.

Hospital de Chancay, Lima, Peru. Oscar Carbajal, M.D.

Hospital Rawson, Cordoba, Argentina. Lorena Ravera M.D.

Hospital Felcio Rocho, Belo Horizonte, Brazil. Mozar Castro M.D.

Hospital Regional Lambayaque, Chiclayo, Peru. Miguel Villegas-Chiroque, M.D., Halbert Christian Sanchez Carrillo, M.D.

Sanatorio Allende, Cordoba, Argentina. Fernando Oscar Riera, M.D., Aldana Mano, M.D.

Hospital Universitario Dr. Jose Eleuterio Gonzalez, Nuevo Leon, Mexico. Adrian Camacho, M.D.

Santa Casa de Misericordia de Porto Alegre, Porto Alegre, Brazil. Claudio Stadnik M.D.

Hospital Nacional Arzobispo Loayza, Lima, Peru. Jorge Gave, M.D.

Hospital Brasilia, Brasilia, Brazil. Rodrigo Biondi. M.D.

Hospital Santa Rosa, Piura, Peru. Ronal Gamarra Velarde, M.D.

Instituto D'OR de Ensino e Pesquisa, Rio de Janeiro, Brazil. Jose Cerbino Neto, M.D.

Hospital Interzonal Dr Jose Penna Bahia Blanca, Bahia Blanca, Argentina. Juan Ditondo, M.D., Myrna Zuain, M.D.

Hospital Ramos Mejia, Buenos Aires, Argentina. Marcelo H. Losso, M.D., Javier J. Toibaro, M.D.

Sanatorio Ramon Cereijo, Caba, Argentina. Mariano Dolz, M.D.

**Other Contributors**

Companies that helped with protocol in the earlier phase of development as part of the ACTIV working group.

AbbVie, North Chicago, IL. David B. Bharucha, M.D., Ph.D., FACC, William Ferguson, Ph.D.

Bristol Myers-Squibb, New York, NY. Brian Gavin, Ph.D., Jonathan Sadeh, M.D. M.Sc., Sheila Kelly, M.D., Michael Maldonado, M.D.

Janssen Pharmaceuticals/Johnson and Johnson, Beerse, Belgium. Richard Melsheimer, Maria Beumont-Mauviel, M.D., Marita Stevens, M.D.

Gilead, Foster City, CA. Huyen Cao, M.D., Adam DeZure, M.D., Kavita Juneja, M.D., Mazin Abdelghany, M.D.

Duke Clinical Research Institute (DCRI), Durham, N.C. Daniel Benjamin, MD, MPH, PhD, Brian Smith, MD, MPH, MHS, Theresa Jasion, Rachel Olson, RN, MS, MBA, Megan Roebuck, M.S., Jacqueline Huvane, Ph.D., Steve McNulty, Pearl Zakroysky, Jun Wen, Adam Silverstein, Jeff Leimberger, Eric Yow, Zhen Huang, Hwasoon Kim, Carla Anderson, Carrie Elliott, RN, Merri Swartz RN, Rose Beci, Susan Halabi, Jyotsna Garg, Hussein Al-Khalidi, Sean O’Brien, PhD

Technical Resources International, Inc (TRI), Bethesda, M.D. Sandra Butler, Ph.D., Daniel Molina, PharmD, MBA; Neta Nelson, MPH, Divya Kalaria, MsCRA, MsFM, Sandhya Rao, MD, M.Ch, Ketty Philogene, MD, Tim Schulz, Ph.D; Averie Kuek, Fatou Bah, MSHS; Jarrard Mitchell, MBA; Elizabeth Polo, M.D., Michelle Wong, J.D.

Syneos Health, Morrisville, NC. Sharon Baldan, Sandra Mendez, M.D., Bradford Stevens, Marcela Toledo, Talita Abba, Emma Herrejon, R.Ph; Cristina Gomez

Fisher Clinical Services, Rockville, MD. Georgeta Mardari, MS, MBA; Neeraja Putta, R.Ph, MBA

University of North Carolina, Chapel Hill, NC – Kevin Anstrom, PhD Lisa LaVange, PhD

Health and Human Services, Washington, D.C. (, Thomas Stock, D.O., William Erhardt, M.D. (H-CORE, formally Operation Warp Speed), Sarah Read, M.D., Mike Proschan, Ph.D. (National Institute of Allergy and Infectious Diseases)

Foundation of the National Institutes of Health, North Bethesda, MD. Stacey Adam, Ph.D.

Biomedical Advanced Research and Development Authority, Washington, D.C. Robin Mason, MS, MBA, Holli Hamilton, M.D.; Derek Eisnor, M.D., Anna O’Rourke, MS, Aditi Patel, Ph.D., RAC, Betty Brody, Anna Chiang, MSW, MPA,

National Center for Advancing Translational Sciences, Bethesda, M.D. Sam Bozzette, M.D., Ph.D., Soju Chang, M.D., MPH, Cynthia Boucher, MS, Jessica Springer, RN, MPH/MPA, Jane Atkinson, D.D.S., Brian Lind, Lilli M. Portilla, MPA, Ami D. Gadhia, JD, LLM, CLP, Sury Vepa, Ph.D., JD, Emily Carlson Marti, MA, Bobbi Gardner, Joni Rutter, Ph.D., Clare Schmitt, Ph.D., Michael Kurilla, M.D.

### **Supplemental Methods**

**Eligibility**

In short, male and non-pregnant females’ ≥18 years of age were eligible if they had laboratory confirmed SARS-CoV-2 infection within 14 days and an anticipated hospital stay of ≥72 hours. Participants were required to have radiological evidence of pulmonary involvement or a new oxygen requirement. Potential candidates were excluded if they had elevated liver function tests (AST or ALT >10 time the upper limit of normal), chronic liver disease, acute kidney injury (estimated glomerular filtration rate <30 ml/min, stable chronic renal insufficiency was permitted), history of New York Heart Association class III/IV congestive heart failure (new right heart failure from COVID-19 was allowed), severe neutropenia (<1.0 x 10^3^/µl) or lymphopenia (<0.2 x 10^3^/µl), or known untreated infection other than COVID-19. Participants were excluded who received cytotoxic or biologic targeted immune-modulator treatments (such as anti-interleukin-1, anti-IL-6, anti-IL-17, or T-cell or B-cell targeted therapies, JAK inhibitors, TNF inhibitors, interferon, or prednisone >10mg per day for more than one month) within 4 weeks or 5 half-lives prior to screening. Vaccination status did not affect eligibility.

**Procedures**

The participant’s clinical status and safety data were captured daily while hospitalized through day 29. If a participant was discharged early, clinical status was collected on day 8, 11, 15, and 29, through an in-person clinic visit or by phone if an in-person visit was not possible. A final safety assessment occurred on day 60 by phone.

Abatacept was administered on day 1 as a single-dose intravenous infusion of 10 mg/kg (maximum dose 1000mg) or matching placebo saline infusion. Remdesivir was given as an intravenous loading dose of 200mg followed by a maintenance dose of 100mg up to 10 days for participants with an estimated glomerular filtration rate>30ml/min. All participants received standard supportive care at the trial site hospital. Dexamethasone treatment was permitted for participants requiring supplemental oxygen or mechanical ventilation in accordance with national guidelines. Emergency use authorized (EUA) monoclonal antibody or COVID-19 convalescent plasma were accepted as SoC treatment. The trial prohibited use of other immunomodulators at the time of randomization. The institutional review board at each site or a centralized institutional review board where applicable approved the trial protocol. The trial was overseen by an independent data and safety monitoring board. Written informed consent was obtained from each participant or from the participant’s legally authorized representative if the participant was unable to provide consent.

**Endpoints**

The primary outcome of time to recovery was evaluated by day 29 by an 8-point ordinal scale (see **Table 1** for definitions). Recovery was identified as the first day a participant attained category 6, 7 or 8. Key secondary end points were pre-specified as 28-day mortality and clinical status at day 14 assessed by ordinal scale improvement from baseline.

**Statistical Analysis**

The sample size requirements were based on the ability to detect a moderate improvement in time to recovery. A total of 788 recoveries are required to provide approximately 85% power to detect a recovery rate ratio (RRR) of 1.25 for abatacept versus placebo, assuming 73% of participants achieve recovery within 28 days, consistent with the ACTT-1 results. The primary efficacy analysis for the abatacept versus shared placebo was based on the Fine-Gray model. Geographic region, age, sex, and baseline 8-point ordinal scale were included as covariates. Three interim analysis were planned to assess futility, and early stopping for efficacy, but due to more rapid accrual than expected, only two were conducted. The Data and Safety Monitoring Board (DSMB) reviewed the results of the two interim analysis.

The statistical analysis plan describes protection for Type 1 error for the primary endpoint and the two key secondary endpoints (28-day mortality and day 14 clinical status) using multiple testing procedure. All-cause mortality through day 28 was analyzed as a binary endpoint with an indicator variable for treatment group through logistic regression. Clinical status through day 14 was calculated using a proportional odds model of improvement on an 8-point ordinal scale by ordinal logistic regression. Adjustments were made for geographic region, baseline 8-point ordinal scale, age, and sex. A multiple imputation approach described in the statistical analysis plan accounted for missing data. Mortality at day 60 and clinical status at day 28 were assessed using similar analyses. Pre-specified subgroup analyses for the primary outcomes evaluated treatment effects across geographic region, duration of symptoms prior to enrollment, baseline disease severity (stratified by variable of moderate and severe, as well as ordinal scale of 4/5 vs 2/3) age, race, sex and comorbidities.

Participants who met eligibility were randomized in a two-stage process. Firstly, each participant was assigned with equal probability to one of the immunomodulatory agents (sub-study) available at the time of enrollment after applying any agent-specific safety exclusions. This step in the randomization was open-label, so participants, clinical providers, and study staff were aware of the sub-study arm. Secondly, each participant was assigned to either the test agent or its matching placebo in an n:1 ratio, where n = the number of agents that the participant was eligible for. The second randomization was blinded. Randomization was stratified by geographic location and disease severity. Moderate disease was defined as hospitalized participants with or without requirement for low-flow supplementary oxygen. Severe disease was defined as hospitalized requiring high flow oxygen devices, non-invasive or mechanical ventilation, or ECMO. In determining which participants in the shared placebo group could be shared for each sub-study, agent specific eligibility criteria were applied. Thus, the shared placebo group utilized for the abatacept analysis was limited to participants who would have been eligible for enrollment in the abatacept sub-study.

All analyses presented in here were based on the modified intention-to-treat (mITT) population, consisting of randomized participants who received at least one dose of the study drug administered in the randomization arm. This population corresponds to the modified intention-to-treat (mITT) cohort in the protocol and statistical analysis plan. The difference between the two populations is 30 individuals: 27 participants who did not receive treatment, and 3 who received the incorrect treatment. The 3 individuals who received the incorrect treatment were excluded because they did not meet the protocol definition for inclusion in the mITT population.

**Safety Assessments**

Safety assessments included a composite endpoint of death, serious adverse events (SAEs), grade 3 (severe) and 4 (potentially life-threatening) adverse events (AEs) occurring through day 60. Secondary infections/superinfections as adverse events of special interest (AESIs) through day 60 and discontinuation or temporary suspension of trial-product administration for any reason were included in the safety assessment. Related SAEs were reviewed in a timely manner by the study safety review team and DSMB.

### **Table S1. Abatacept vs. shared placebo**

| **Endpoint** | **Efficacy boundary -**  **t-statistic and p-value** | **Observed data -**  **t-statistic and nominal p-value** | **Decision based on the gatekeeper approach** |
| --- | --- | --- | --- |
| Time-to-recovery | t>1.9869  p<0.0469 | t=1.9002  p=0.0574 | Fail to reject the null hypothesis and stop the hierarchical testing process |
| 28-day mortality endpoint | t<-2.5624  p<0.0104 | t=-2.2600  p=0.0238 |  |
| 14-day 8-point ordinal scale | t>2.2839  p<0.0224 | t=1.4630  p=0.1435 |  |

| **Interim Analysis** | **Information Fraction** |
| --- | --- |
| First Interim Analysis | 0.3109 |
| Second Interim Analysis | 0.6345 |
| Final Analysis | 1.0000 |

### **Table S2. Demographics and baseline characteristics – US (modified intent-to-treat)**

| **Demographics and baseline characteristics** | **Abatacept**    **N=340** | **Placebo**    **N=342** | **All Subjects**    **N=682** |
| --- | --- | --- | --- |
| Age in years* | 55.4 (14.42) | 56.8 (14.47) | 56.1 (14.45) |
| Male sex | 220 (64.7%) | 186 (54.4%) | 406 (59.5%) |
| Race |  |  |  |
| White | 214 (62.9%) | 220 (64.3%) | 434 (63.6%) |
| Black or African American | 66 (19.4%) | 67 (19.6%) | 133 (19.5%) |
| American Indian or Alaska Native | 1 (0.3%) | 4 (1.2%) | 5 (0.7%) |
| Asian | 16 (4.7%) | 16 (4.7%) | 32 (4.7%) |
| Native Hawaiian or Other Pacific Islander | 1 (0.3%) | 0 (0.0%) | 1 (0.1%) |
| Multiracial | 0 (0.0%) | 2 (0.6%) | 2 (0.3%) |
| Other | 14 (4.1%) | 8 (2.3%) | 22 (3.2%) |
| Unknown | 28 (8.2%) | 25 (7.3%) | 53 (7.8%) |
| Hispanic or Latino Ethnicity | 56 (16.5%) | 78 (22.8%) | 134 (19.6%) |
| Disease severity at baseline^†^ |  |  |  |
| Severe disease | 142 (41.8%) | 141 (41.2%) | 283 (41.5%) |
| Moderate disease | 198 (57.2%) | 201 (58.8%) | 399 (58.5%) |
| Clinical status (8-point ordinal scale) at baseline |  |  |  |
| 1. Death | 0 | 0 | 0 |
| 2. Hospitalized, on invasive ventilation or ECMO | 29 (8.5%) | 27 (7.9%) | 56 (8.2%) |
| 3. Hospitalized, on non-invasive ventilation or high flow oxygen devices | 113 (33.2%) | 114 (33.3%) | 227 (33.3%) |
| 4. Hospitalized, requiring supplemental oxygen | 179 (52.6%) | 188 (55.0%) | 367 (53.8%) |
| 5. Hospitalized, not requiring supplement oxygen, requiring ongoing medical care | 19 (5.6%) | 13 (3.8%) | 32 (4.7%) |
| 6. Hospitalized, not requiring supplement oxygen, not requiring ongoing medical care | 0 | 0 | 0 |
| 7. Not hospitalized, limitations on activity and/or requiring home oxygen | 0 | 0 | 0 |
| 8. Not hospitalized, no limitations on activities | 0 | 0 | 0 |
| Geographic Region |  |  |  |
| USA - Northeast | 124 (36.5%) | 126 (36.8%) | 250 (36.7%) |
| USA - Midwest | 82 (24.1%) | 77 (22.5%) | 159 (23.3%) |
| USA - South | 96 (28.2%) | 96 (28.1%) | 192 (28.2%) |
| USA - West | 38 (11.2%) | 43 (12.6%) | 81 (11.9%) |
| Days from Symptom Onset* | -8.6 (4.49) | -9.3 (6.23) | -9.0 (5.44) |
| Body Mass Index (kg/m^2^)* | 33.6 (9.15) | 34.2 (9.06) | 33.9 (9.10) |
| **Comorbidities** |  |  |  |
| Hypertension | 155 (45.6%) | 165 (48.2%) | 320 (46.9%) |
| Obesity (BMI>=30 kg/m^2^) | 199 / 334 (59.6%) | 216 / 329 (65.7%) | 415 / 663 (62.6%) |
| Diabetes Mellitus | 113 (33.2%) | 115 (33.6%) | 228 (33.4%) |
| Coronary Artery Disease | 28 (8.2%) | 26 (7.6%) | 54 (7.9%) |
| History of Heart Failure | 14 (4.1%) | 13 (3.8%) | 27 (4.0%) |
| History of Cancer | 29 (8.5%) | 32 (9.4%) | 61 (8.9%) |
| Asthma | 35 (10.3%) | 44 (12.9%) | 79 (11.6%) |
| Chronic Obstructive Pulmonary Disease | 15 (4.4%) | 20 (5.8%) | 35 (5.1%) |
| Tuberculosis | 1 (0.3%) | 2 (0.6%) | 3 (0.4%) |
| HIV/AIDS | 1 (0.3%) | 4 (1.2%) | 5 (0.7%) |
| Severe Liver Disease | 1 (0.3%) | 2 (0.6%) | 3 (0.4%) |
| Severe Kidney Disease | 3 (0.9%) | 6 (1.8%) | 9 (1.3%) |

 * Displayed as Mean (Std. Dev.)

^†^Disease severity at baseline calculated as moderate disease = ordinal 5 + ordinal 4, severe disease = ordinal 3 + ordinal 2

### **Table S2 (continued). Demographics and baseline characteristics – Latin America (modified intent-to-treat)**

| **Demographics and baseline characteristics** | **Abatacept**    **N=169** | **Placebo**    **N=168** | **All Subjects**    **N=337** |
| --- | --- | --- | --- |
| Age in years* | 53.2 (14.93) | 51.2 (14.46) | 52.2 (14.71) |
| Male sex | 101 (59.8%) | 109 (64.9%) | 210 (62.3%) |
| Race |  |  |  |
| White | 104 (61.5%) | 102 (60.7%) | 206 (61.1%) |
| Black or African American | 4 (2.4%) | 3 (1.8%) | 7 (2.1%) |
| American Indian or Alaska Native | 4 (2.4%) | 0 (0.0%) | 4 (1.2%) |
| Asian | 0 (0.0%) | 0 (0.0%) | 0 (0.0%) |
| Native Hawaiian or Other Pacific Islander | 0 (0.0%) | 0 (0.0%) | 0 (0.0%) |
| Multiracial | 0 (0.0%) | 1 (0.6%) | 1 (0.3%) |
| Other | 57 (33.7%) | 62 (36.9%) | 119 (35.3%) |
| Unknown | 0 (0.0%) | 0 (0.0%) | 0 (0.0%) |
| Hispanic or Latino Ethnicity | 158 (93.5%) | 158 (94.0%) | 316 (93.8%) |
| Disease severity at baseline^†^ |  |  |  |
| Severe disease | 78 (46.2%) | 81 (48.2%) | 159 (47.2%) |
| Moderate disease | 91 (53.8%) | 87 (51.8%) | 178 (52.8%) |
| Clinical status (8-point ordinal scale) at baseline |  |  |  |
| 1. Death | 0 | 0 | 0 |
| 2. Hospitalized, on invasive ventilation or ECMO | 18 (10.7%) | 24 (14.3%) | 42 (12.5%) |
| 3. Hospitalized, on non-invasive ventilation or high flow oxygen devices | 60 (35.5%) | 57 (33.9%) | 117 (34.7%) |
| 4. Hospitalized, requiring supplemental oxygen | 87 (51.5%) | 82 (48.8%) | 169 (50.1%) |
| 5. Hospitalized, not requiring supplement oxygen, requiring ongoing medical care | 4 (2.4%) | 5 (3.0%) | 9 (2.7%) |
| 6. Hospitalized, not requiring supplement oxygen, not requiring ongoing medical care | 0 | 0 | 0 |
| 7. Not hospitalized, limitations on activity and/or requiring home oxygen | 0 | 0 | 0 |
| 8. Not hospitalized, no limitations on activities | 0 | 0 | 0 |
| Geographic Region |  |  |  |
| Argentina | 52 (30.8%) | 54 (32.1%) | 106 (51.5%) |
| Brazil | 45 (26.6%) | 45 (26.8%) | 90 (26.7%) |
| Mexico | 18 (10.7%) | 16 (9.5%) | 34 (10.1%) |
| Peru | 54 (32.0%) | 53 (31.5%) | 107 (31.8%) |
| Days from Symptom Onset* | -5.3 (3.41) | -6.4 (3.52) | -5.9 (3.50) |
| Body Mass Index (kg/m^2^)* | 30.6 (5.34) | 29.9 (4.98) | 30.2 (5.17) |
| **Comorbidities** |  |  |  |
| Hypertension | 58 (34.3%) | 45 (26.8%) | 103 (30.6%) |
| Obesity (BMI>=30 kg/m^2^) | 88 (52.1%) | 82 (48.8%) | 170 (50.4%) |
| Diabetes Mellitus | 33 (19.5%) | 26 (15.5%) | 59 (17.5%) |
| Coronary Artery Disease | 5 (3.0%) | 3 (1.8%) | 8 (2.4%) |
| History of Heart Failure | 2 (1.2%) | 1 (0.6%) | 3 (0.9%) |
| History of Cancer | 2 (1.2%) | 4 (2.4%) | 6 (1.8%) |
| Asthma | 1 (0.6%) | 9 (5.4%) | 10 (3.0%) |
| Chronic Obstructive Pulmonary Disease | 4 (2.4%) | 4 (2.4%) | 8 (2.4%) |
| Tuberculosis | 2 (1.2%) | 2 (1.2%) | 4 (1.2%) |
| HIV/AIDS | 2 (1.2%) | 0 (0.0%) | 2 (0.6%) |
| Severe Liver Disease | 0 (0.0%) | 0 (0.0%) | 0 (0.0%) |
| Severe Kidney Disease | 1 (0.6%) | 2 (1.2%) | 3 (0.9%) |

 * Displayed as Mean (Std. Dev.)

^†^Disease severity at baseline calculated as moderate disease = ordinal 5 + ordinal 4, severe disease = ordinal 3 + ordinal 2

### **Table S3. Additional secondary endpoints (modified intent-to-treat population)**

|  | **Proportion of Participants** | |  | **Abatacept versus Shared Placebo** | |
| --- | --- | --- | --- | --- | --- |
|  | **Abatacept** | **Shared placebo** | | **OR Estimate (95% CI)** | ***p* value** |
| **Mortality^*^ at day 14** | 24/509 (4.7%) | 40/510 (7.8%) | | 0.552 (0.317, 0.963) | 0.0363 |
| **Mortality at Day 28** | 56/509 (11.0%) | 77/510 (15.1%) | | 0.622 (0.411, 0.939) | 0.0238 |
| **Clinical status^†^ at day 14 (N)** | 489 | 494 | | 1.190 (0.943, 1.503) | 0.1435 |
| **Clinical status at day 28 (N)** | 483 | 488 | | 1.353 (1.055, 1.734) | 0.0172 |

^*^Mortality at 14 and 28 days calculated as odds of dying using logistic regression. A number less than 1 favors Abatacept.

^†^Clinical status at 14 and 28 days calculated as proportional odds model by ordinal logistic regression. A number greater than 1 favors Abatacept.

### **Table S4. Demographics and baseline characteristics (intent-to-treat)**

| **Demographics and baseline characteristics** | **Abatacept**    **N=524** | **Placebo**    **N=525** | **All Subjects**    **N=1049** |
| --- | --- | --- | --- |
| Age in years* | 54.8 (14.65) | 55.0 (14.66) | 54.9 (14.65) |
| Male sex | 327 (62.4%) | 303 (57.7%) | 630 (60.1%) |
| Race |  |  |  |
| White | 325 (62.0%) | 327 (62.3%) | 652 (62.2%) |
| Black or African American | 74 (14.1%) | 76 (14.5%) | 150 (14.3%) |
| American Indian or Alaska Native | 5 (1.0%) | 5 (1.0%) | 10 (1.0%) |
| Asian | 16 (3.1%) | 17 (3.2%) | 33 (3.1%) |
| Native Hawaiian or Other Pacific Islander | 1 (0.2%) | 0 (0.0%) | 1 (0.1%) |
| Multiracial | 0 (0.0%) | 3 (0.6%) | 3 (0.3%) |
| Other | 74 (14.1%) | 71 (13.5%) | 145 (13.8%) |
| Unknown | 29 (5.5%) | 26 (5.0%) | 55 (5.2%) |
| Hispanic or Latino Ethnicity | 219 (41.8%) | 242 (46.1%) | 461 (43.9%) |
| Disease severity at baseline^†^ |  |  |  |
| Severe disease | 223 (42.6%) | 226 (43.0%) | 449 (42.8%) |
| Moderate disease | 301 (57.4%) | 299 (57.0%) | 600 (57.2%) |
| Clinical status (8-point ordinal scale) at baseline |  |  |  |
| 1. Death | 0 | 0 | 0 |
| 2. Hospitalized, on invasive ventilation or ECMO | 48 (9.2%) | 51 (9.7%) | 99 (9.4%) |
| 3. Hospitalized, on non-invasive ventilation or high flow oxygen devices | 175 (33.4%) | 175 (33.3%) | 350 (33.4%) |
| 4. Hospitalized, requiring supplemental oxygen | 276 (52.7%) | 279 (53.1%) | 555 (52.9%) |
| 5. Hospitalized, not requiring supplement oxygen, requiring ongoing medical care | 25 (4.8%) | 20 (3.8%) | 45 (4.3%) |
| 6. Hospitalized, not requiring supplement oxygen, not requiring ongoing medical care | 0 | 0 | 0 |
| 7. Not hospitalized, limitations on activity and/or requiring home oxygen | 0 | 0 | 0 |
| 8. Not hospitalized, no limitations on activities | 0 | 0 | 0 |
| Geographic Region |  |  |  |
| Argentina | 52 (9.9%) | 57 (10.9%) | 109 (10.4%) |
| Brazil | 46 (8.8%) | 45 (8.6%) | 91 (8.7%) |
| Mexico | 18 (3.4%) | 16 (3.0%) | 34 (3.2%) |
| Peru | 56 (10.7%) | 55 (10.5%) | 111 (10.6%) |
| USA - Northeast | 130 (24.8%) | 130 (24.8%) | 260 (24.8%) |
| USA - Midwest | 83 (15.8%) | 80 (15.2%) | 163 (15.5%) |
| USA - South | 100 (19.1%) | 99 (18.9%) | 199 (19.0%) |
| USA - West | 39 (7.4%) | 43 (8.2%) | 82 (7.8%) |
| Days from Symptom Onset* | -9.3 (4.33) | -9.9 (5.53) | -9.6 (4.98) |
| Body Mass Index (kg/m^2^)* | 32.6 (8.23) | 32.7 (8.16) | 32.7 (8.19) |
| **Comorbidities** |  |  |  |
| Hypertension | 219 (41.8%) | 218 (41.5%) | 437 (41.7%) |
| Obesity (BMI>=30 kg/m^2^) | 297 / 518 (57.3%) | 308 / 512 (60.2%) | 605 / 1030 (58.7%) |
| Diabetes Mellitus | 149 (28.4%) | 146 (27.8%) | 295 (28.1%) |
| Coronary Artery Disease | 34 (6.5%) | 30 (5.7%) | 64 (6.1%) |
| History of Heart Failure | 16 (3.1%) | 15 (2.9%) | 31 (3.0%) |
| History of Cancer | 31 (5.9%) | 36 (6.9%) | 67 (6.4%) |
| Asthma | 38 (7.3%) | 55 (10.5%) | 93 (8.9%) |
| Chronic Obstructive Pulmonary Disease | 20 (3.8%) | 24 (4.6%) | 44 (4.2%) |
| Tuberculosis | 3 (0.6%) | 4 (0.8%) | 7 (0.7%) |
| HIV/AIDS | 3 (0.6%) | 5 (1.0%) | 8 (0.8%) |
| Severe Liver Disease | 1 (0.2%) | 2 (0.4%) | 3 (0.3%) |
| Severe Kidney Disease | 4 (0.8%) | 9 (1.7%) | 13 (1.2%) |
| **Concomitant medication** |  |  |  |
| Remdesivir (Day 1 - Day 5) | 488 (93.1%) | 493 (93.9%) | 981 (93.5%) |
| Corticosteroids (Day 1 - Day 5) | 468 (89.3%) | 488 (93.0%) | 956 (91.1%) |
| Tocilizumab (any time post randomization) | 14 (2.7%) | 15 (2.9%) | 29 (2.8%) |
| Baricitinib (any time post randomization) | 6 (1.1%) | 14 (2.7%) | 20 (1.9%) |

 * Displayed as Mean (Std. Dev.)

^†^Disease severity at baseline calculated as moderate disease = ordinal 5 + ordinal 4, severe disease = ordinal 3 + ordinal 2

### **Table S5. Primary and secondary endpoints (intent-to-treat population)**

|  | **Proportion of participants** | |  | **Abatacept versus Shared Placebo** | |
| --- | --- | --- | --- | --- | --- |
|  | **Abatacept** | **Placebo** | | **Estimate (95% CI)** | ***p* value** |
| **Recovery through day 28** | 414/524 (79.0%) | 397/525 (75.6%) | | 1.120 (0.984, 1.275) | 0.0864 |
| **Mortality at day 14** | 24/524 (4.6%) | 40/525 (7.6%) | | 0.552 (0.317, 0.960) | 0.0354 |
| **Mortality at day 28** | 56/524 (10.7%) | 77/525 (14.7%) | | 0.629 (0.416, 0.951) | 0.0281 |
| **Clinical status at day 14 (N)** | 493 | 499 | | 1.174 (0.932, 1.480) | 0.1732 |
| **Clinical status at day 28 (N)** | 487 | 493 | | 1.338 (1.041, 1.718) | 0.0228 |

Estimates represent Recovery rate ratio, RRR (time to recovery) and Odds ratio, OR (mortality and clinical status)

### **Figure S1. Consort diagram for abatacept substudy of ACTIV-1 IM master protocol**

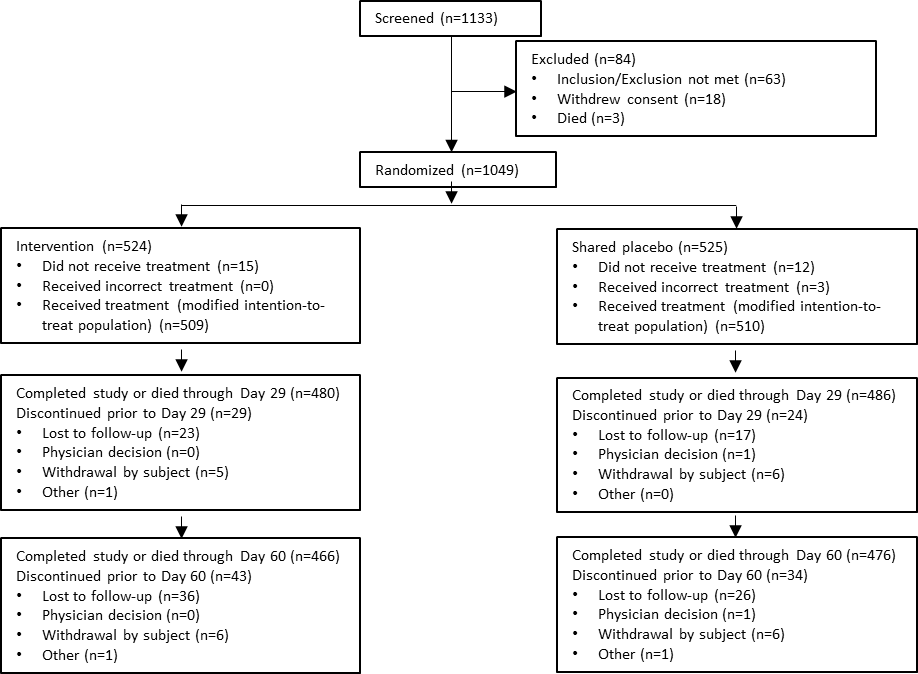

### **Figure S2. Forest plot of recovery rate ratios of time to recovery through day 28 by subgroup (modified intent-to-treat population)**

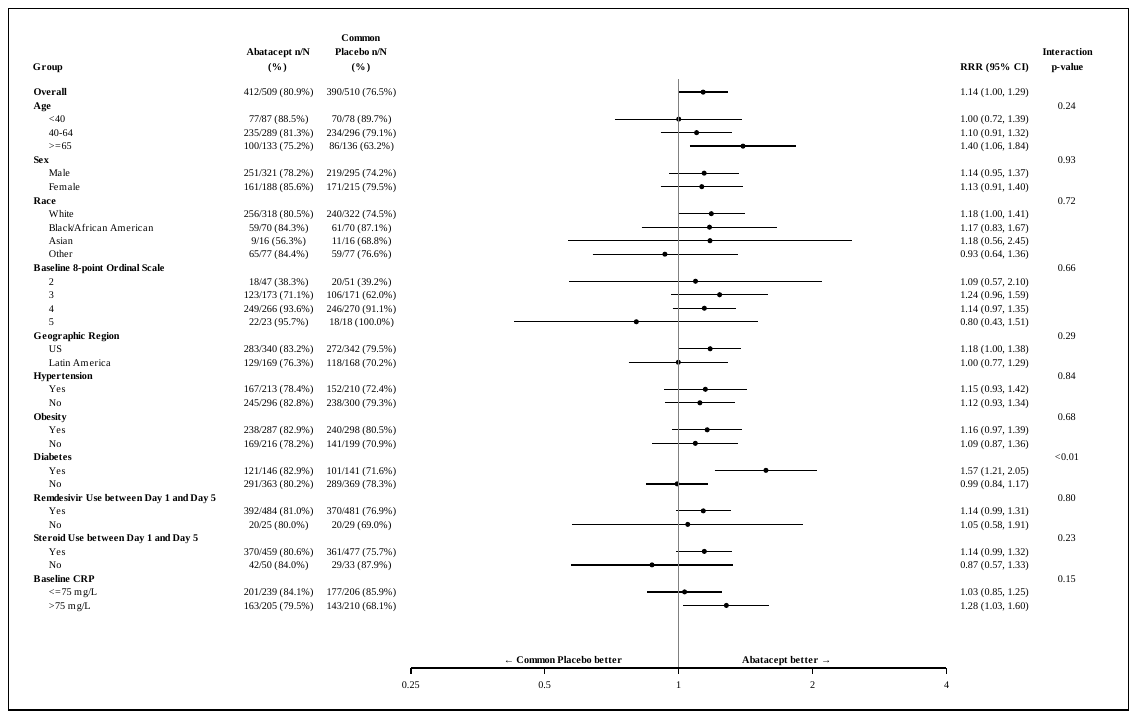

### **Figure S3. Forest plot of proportional odds model of clinical status score at day 14 by subgroup (modified intent-to-treat population)**

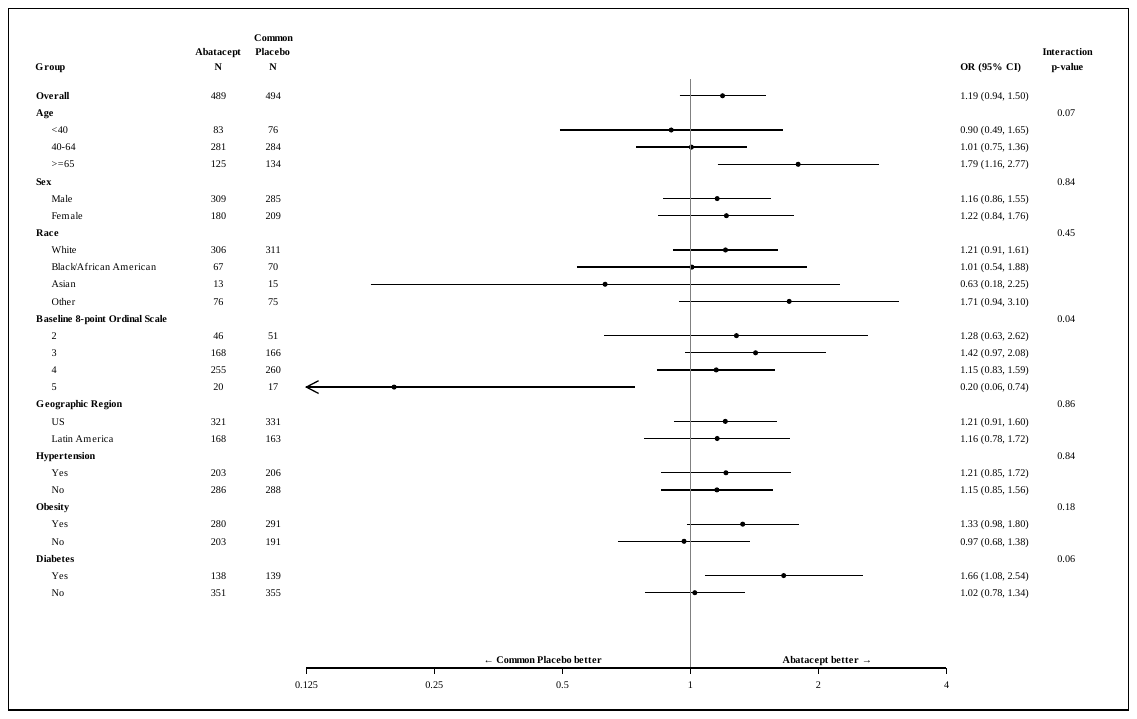
